# Delivering maternal and child health interventions through the private sector in LMIC: a scoping review of strategies and effective approaches

**DOI:** 10.1101/2023.03.17.23287397

**Authors:** Phyllis Awor

## Abstract

Despite growing evidence on the role of private health providers, the global public health response to date has primarily focused on the provision of public sector health services for women and children in low and middle income countries (LMIC). Limitations of this approach are well documented and include: shortage of human resources, inefficient institutional frameworks and inadequate quality especially in rural areas. In order to achieve Universal Health Coverage, it is important to strengthen both the public and private sector. The objective was to determine effective strategies for engagement with private health providers for maternal and child health in LMIC.

A scoping review of both published and grey literature from 2000 – 2022 was undertaken, using including all types of papers reporting on: either population level data on the extent of utilization of the private sector for maternal and child health services in more than one country; or interventions for service provision in the private sector using population level results in one or more country; Or both of the above. Aggregate results were extracted, and content analysis was used to identify engagement strategies/themes.

The results confirm that the private sector is the dominant provider of outpatient care for women and children in LMIC, and a significant provider of reproductive and maternal health services including for inpatient care. Effective strategies and recommendations for engagement with private health providers are presented across challenges of private health provider engagement.

## INTRODUCTION

While Universal health coverage (UHC) is the aspiration for all governments and primary health care offers a cost-effective route to achieving UHC, most primary health systems are weak in low and middle income countries and they cannot provide the comprehensive, people-centered and integrated care needed (Langlois et al., 2020, WHO, 2018a). In an attempt to meet the health needs of people in LMIC, health systems have evolved and are pluralistic in nature, having various stakeholders and actors working side by side, to provide health services. This often includes the existence of public and private providers as well as allopathic and alternative providers, all within a health system (Bloom et al., 2011, WHO, 2020a).

The private sector particularly plays a significant role in the delivery of health services in LMIC, providing more than half of all health care in sub-Saharan Africa (SSA) and up to three quarters of health care in Asia. Private health providers also deliver about two thirds (60-70%) of treatment for fever, cough and diarrhoea in children; and up to one third (30-40%) of key maternal health services including institutional delivery, antenatal care and modern contraceptives in LMIC (Grepin, 2016, Montagu and Chakraborty, 2021, Campbell et al., 2016).

Despite the increasing evidence of the role of private providers in delivery of maternal and child health services in LMIC, the global public health response to date has primarily focused on the provision of public sector health services for women, infants and children. And yet the health systems are struggling with low public financing of health care, high individual out of pocket expenditure and high use of private health care providers [14]. It is evident that in order to achieve Universal Health Coverage in this setting, it is important to strengthen both the public and private sector. Engagement with private health care providers and coordination of health care provision also in the private sector is crucial (Morgan et al., 2016, Awor et al., 2018). This can contribute to strengthened local health systems, using proven and context specific interventions which empower people and communities for better health – the core principles of primary health care. The objective of this scoping review was to therefore to identify effective strategies for engagement with private health providers for maternal and child health service provision at the primary health care level.

### Definitions

Private sector: The private sector is very heterogeneous and here we define it as “those individuals or organizations providing health services or products that are not directly owned or controlled by government” (WHO, 2018b). The private sector can be classified into the following diverse categories, each having different attributes and purposes: for-profit and not-for-profit; formal and informal; and even domestic and foreign. The private sector health services are often categorized into outpatient and inpatient services. Notably, outpatient services are provided in clinic settings as well as at drug dispensing outlets including pharmacies and drug shops.

Non state (private) providers: “All providers who exist outside the public sector whether their aim is philanthropic or commercial and whose aim is to treat illness or prevent disease. They include: large and small commercial companies, groups of professionals like doctors, national and international NGOs and individual providers and shop keepers. The services they provide include hospital, nursing and maternity homes, clinics run by doctors, nurses, midwives and paramedical workers, diagnostic facilities, like labs and radiological units, and the sale of drugs from pharmacies and unqualified static and itinerant drug sellers including general stores.” (Mills et al., 2002)

Private sector engagement refers to a partnership between the public and private sectors to achieve a specified goal (WHO, 2018b).

## METHODS

A scoping literature review was undertaken, with the following objectives:

1. To identify the extent of utilization of private health providers for primary health care by women and children in low and middle income countries.
2. To determine effective strategies for engagement with private health providers for maternal and child health primary health care service provision.

The literature search was conducted using both PubMed and Google scholar including literature from 2000-2020. A combination of the following search terms were used to conduct the literature search, up to May 2022 – “Private sector”, “engagement” , “public-private partnership”, “maternal health”, “child health”, “primary health care”. All types of studies were included provided that they reported on:

a. Either population level data on the extent of utilization of the private sector for maternal (including delivery) and child health services in more than one country;
b. Or interventions (including public-private engagement) for service provision in the private sector using population level results in one or more country;
c. Or both of the above.

### Handling of the search results and identified papers

References were exported using the “CSV” option into excel and endnote. The lead author and a research assistant independently reviewed all the titles of the identified articles and selected those most appropriate for the search. Any discrepancies were resolved by the lead author. All abstracts were then screened by both researchers and the final papers agreed upon for inclusion. The full text of each of the selected papers was reviewed and data was extracted into a pre-designed data extraction sheet in excel.

### Analysis

for objective 1 on the extent of utilization of the private sector for maternal and child health, the aggregate results were extracted and are presented in table 1. For objective 2 on effective strategies and interventions for private provider engagement, quantitative data is extracted as results; and thematic content analysis was used to identify key themes emerging from the data and the paper. These were complemented by pre-determined themes (strategies) already identified in the literature, for which data was also extracted. The pre-determined themes included the various private sector engagement strategies, challenges and recommendations for effective engagement. These strategies are presented in table 2 and are discussed further below.

**Table 1:**
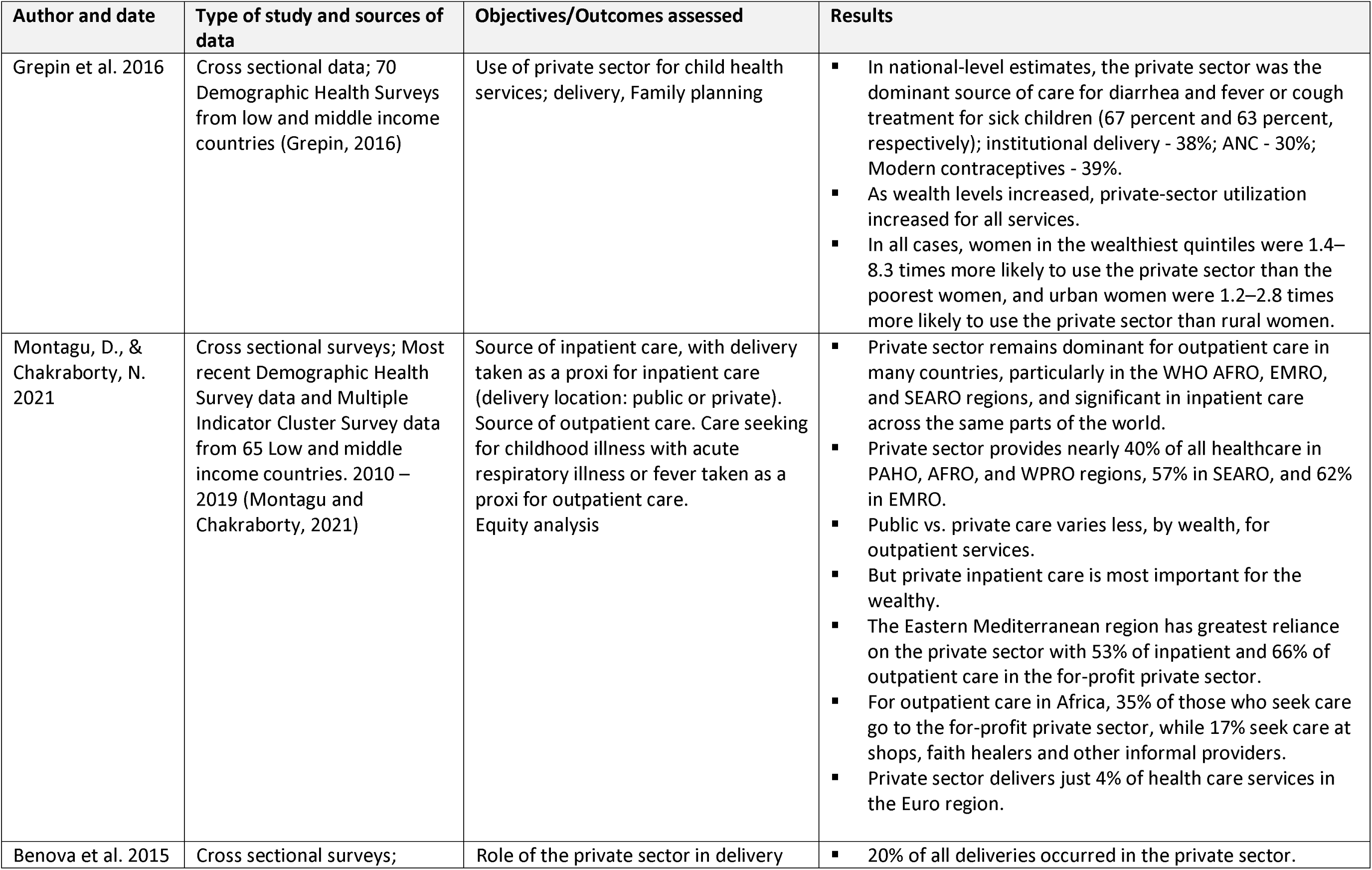
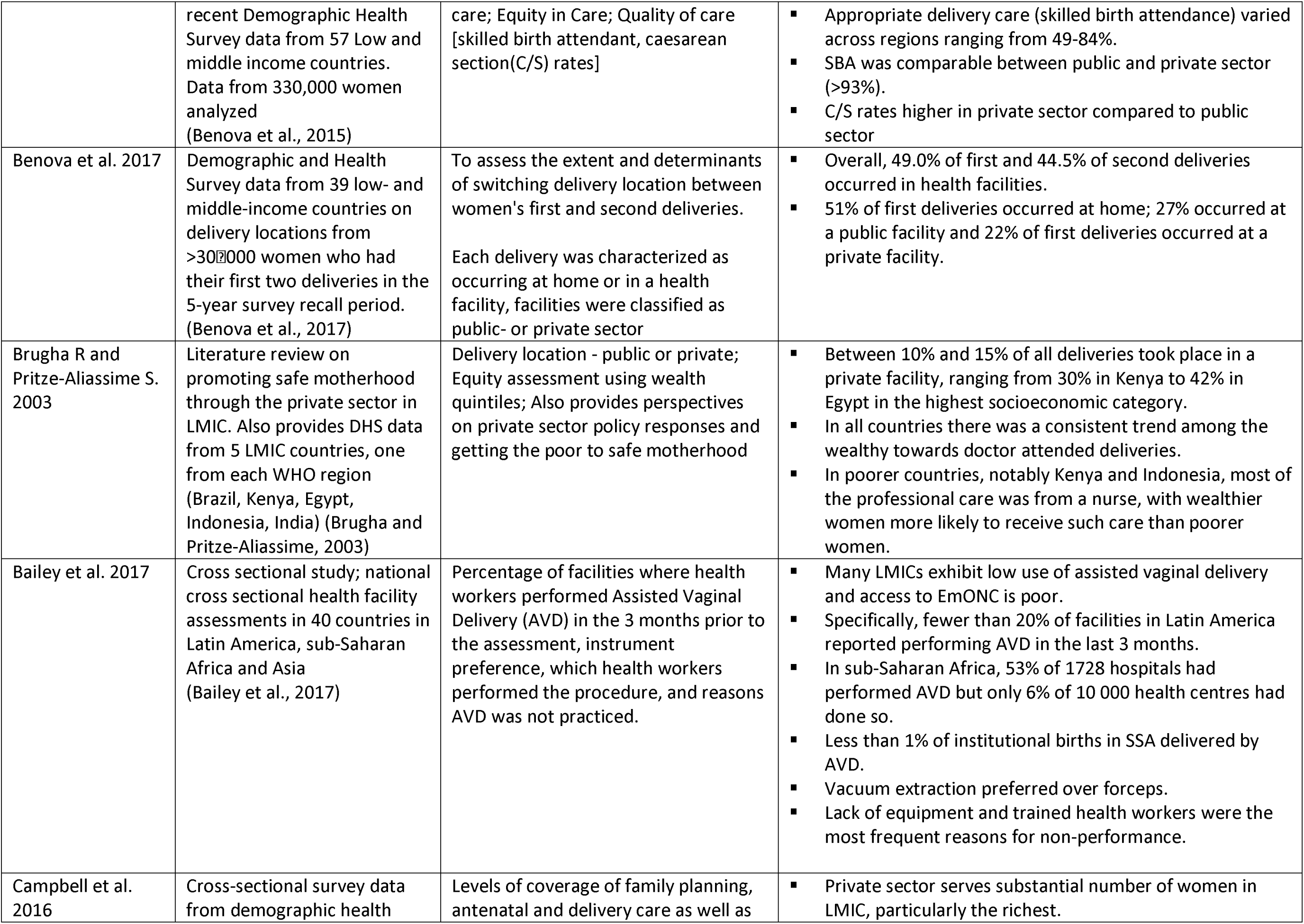
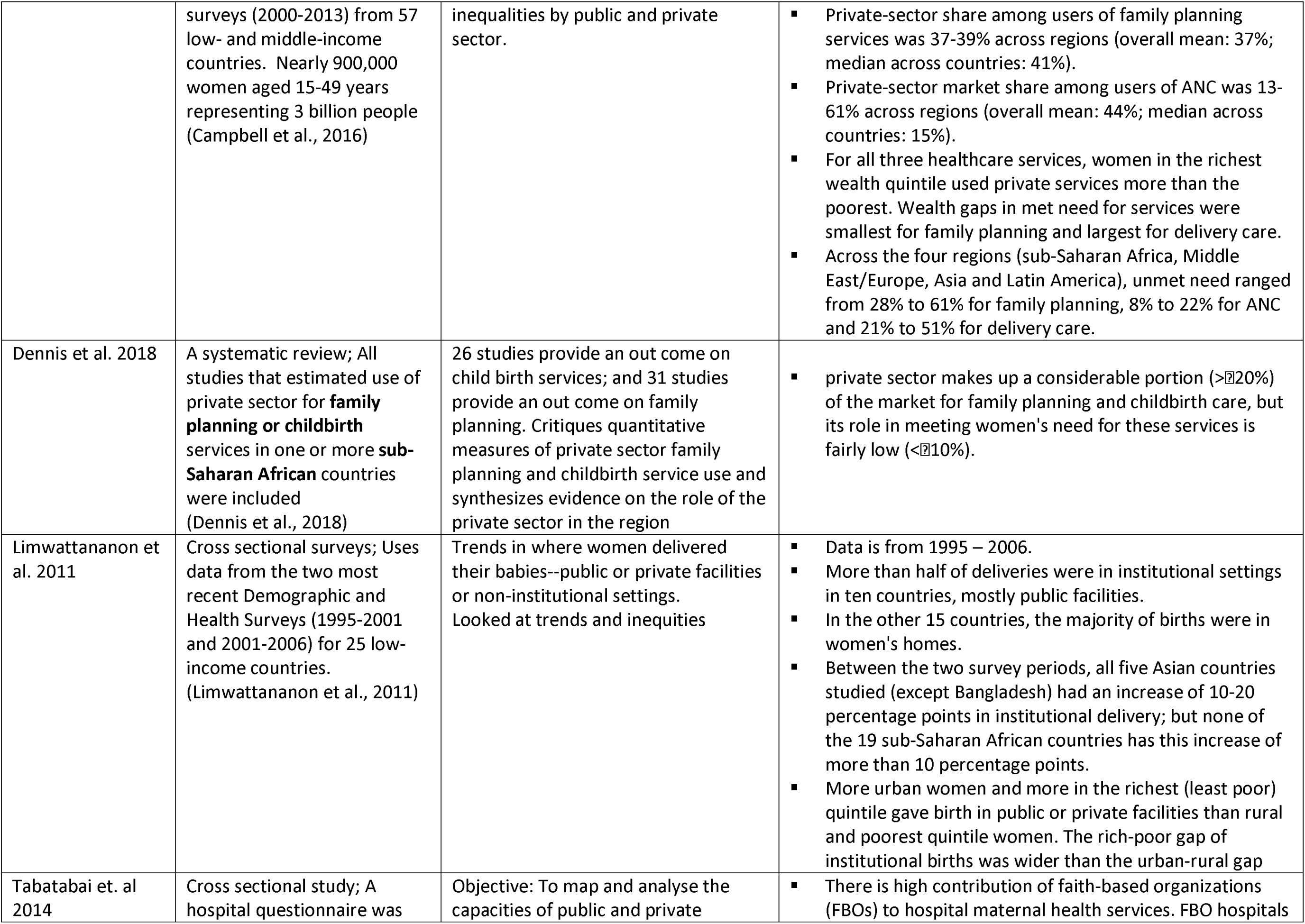
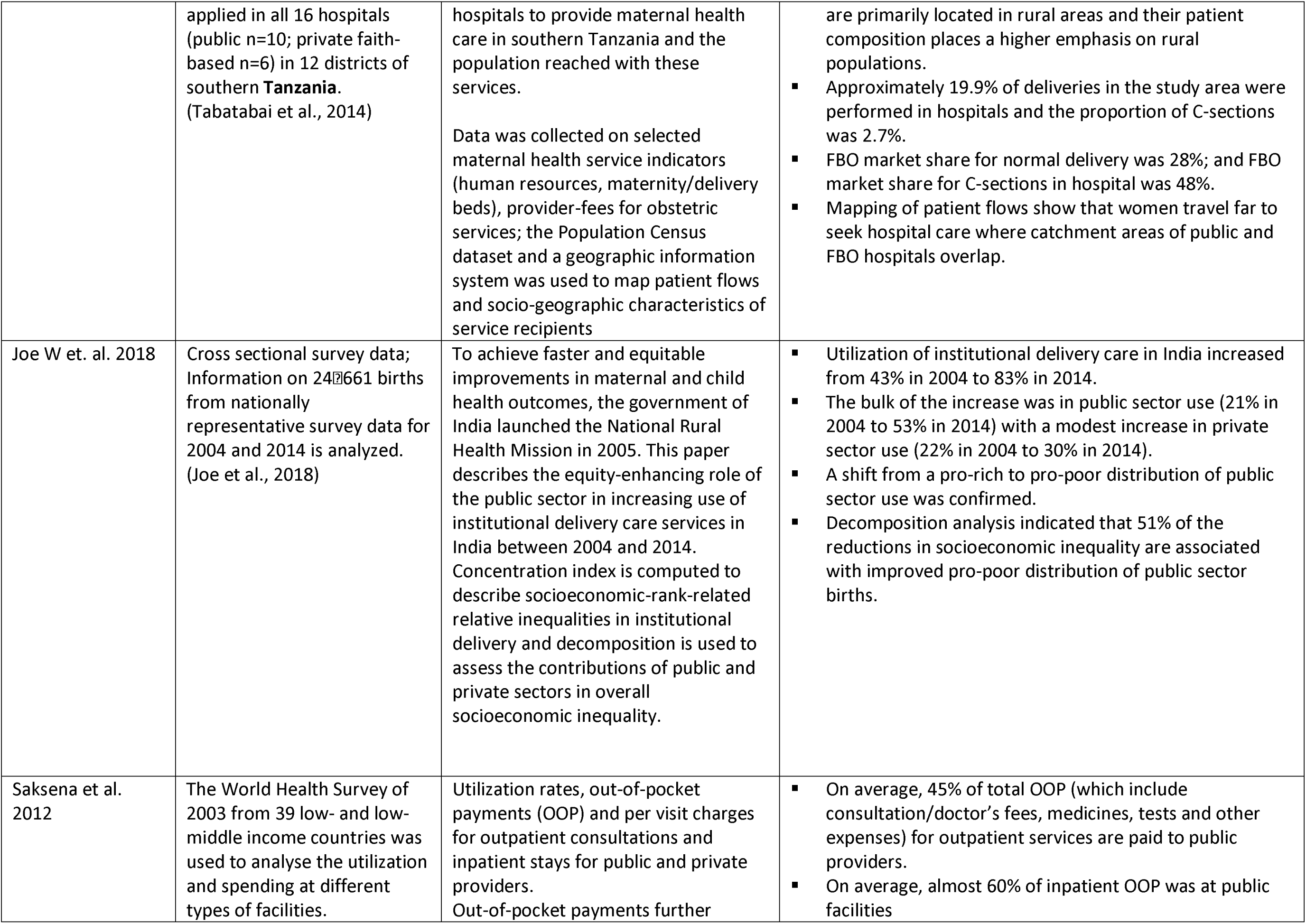
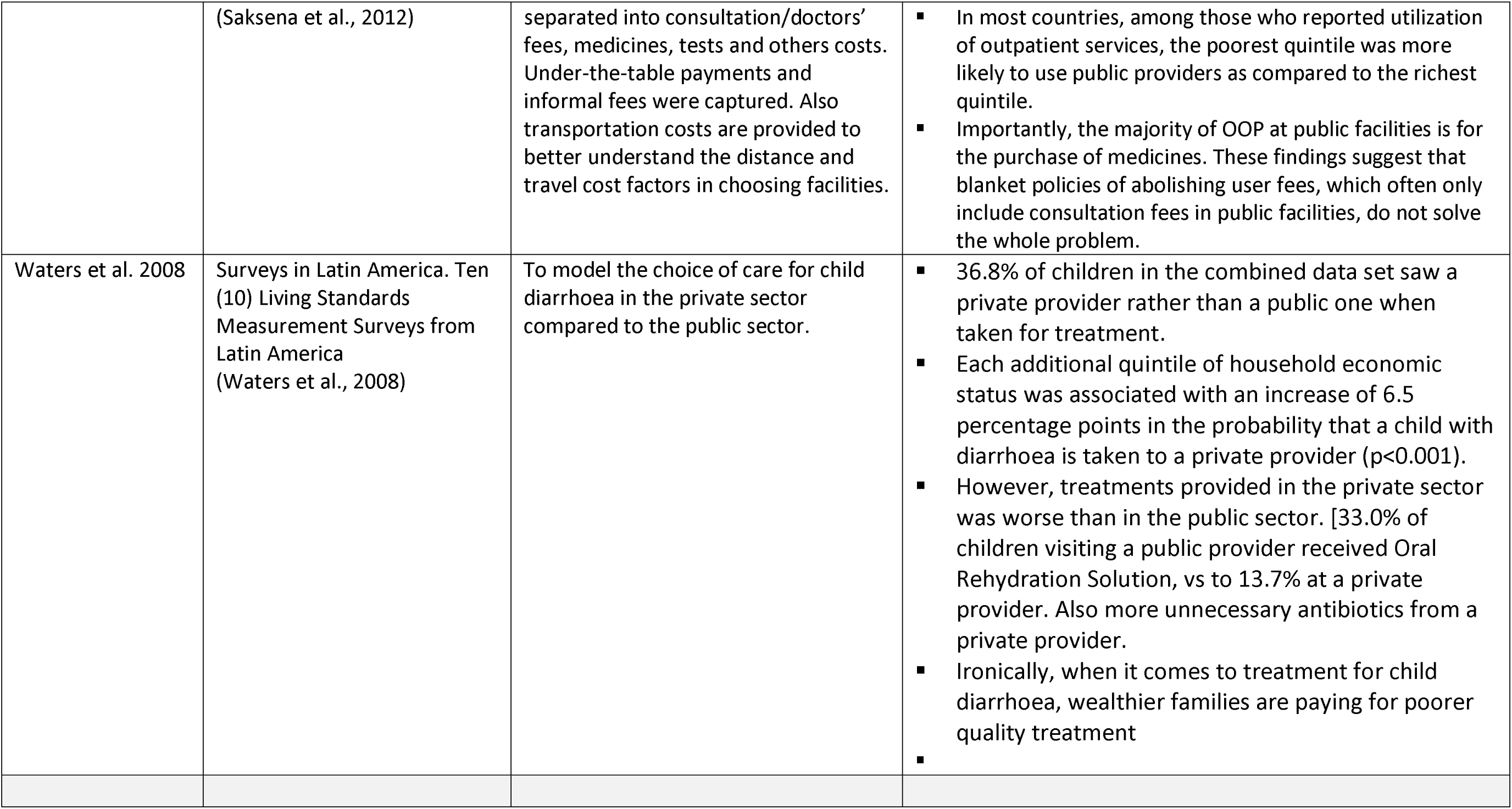
Extent of utilization of private health providers for maternal and child health services in LMIC

**Table 2:**
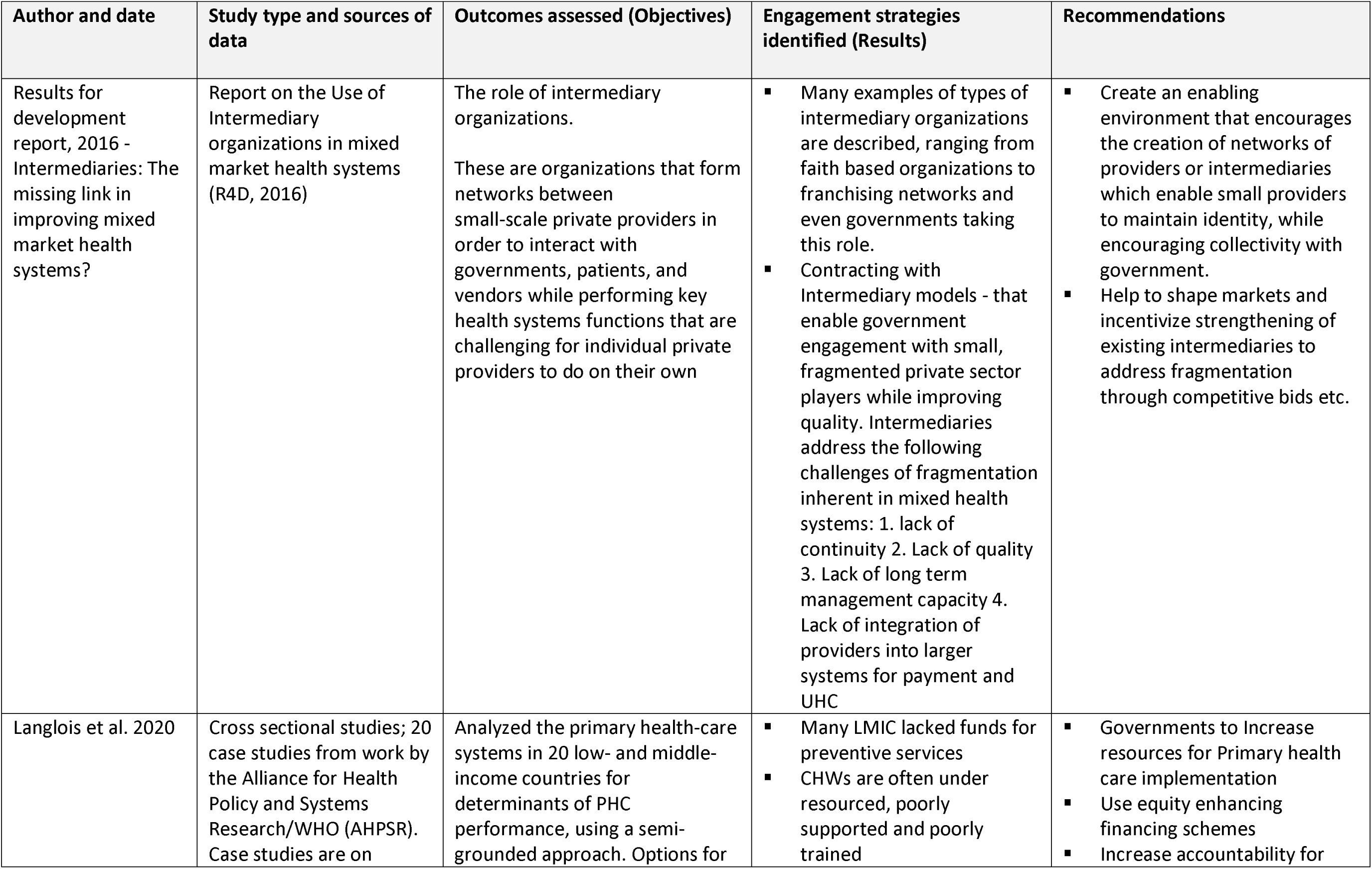
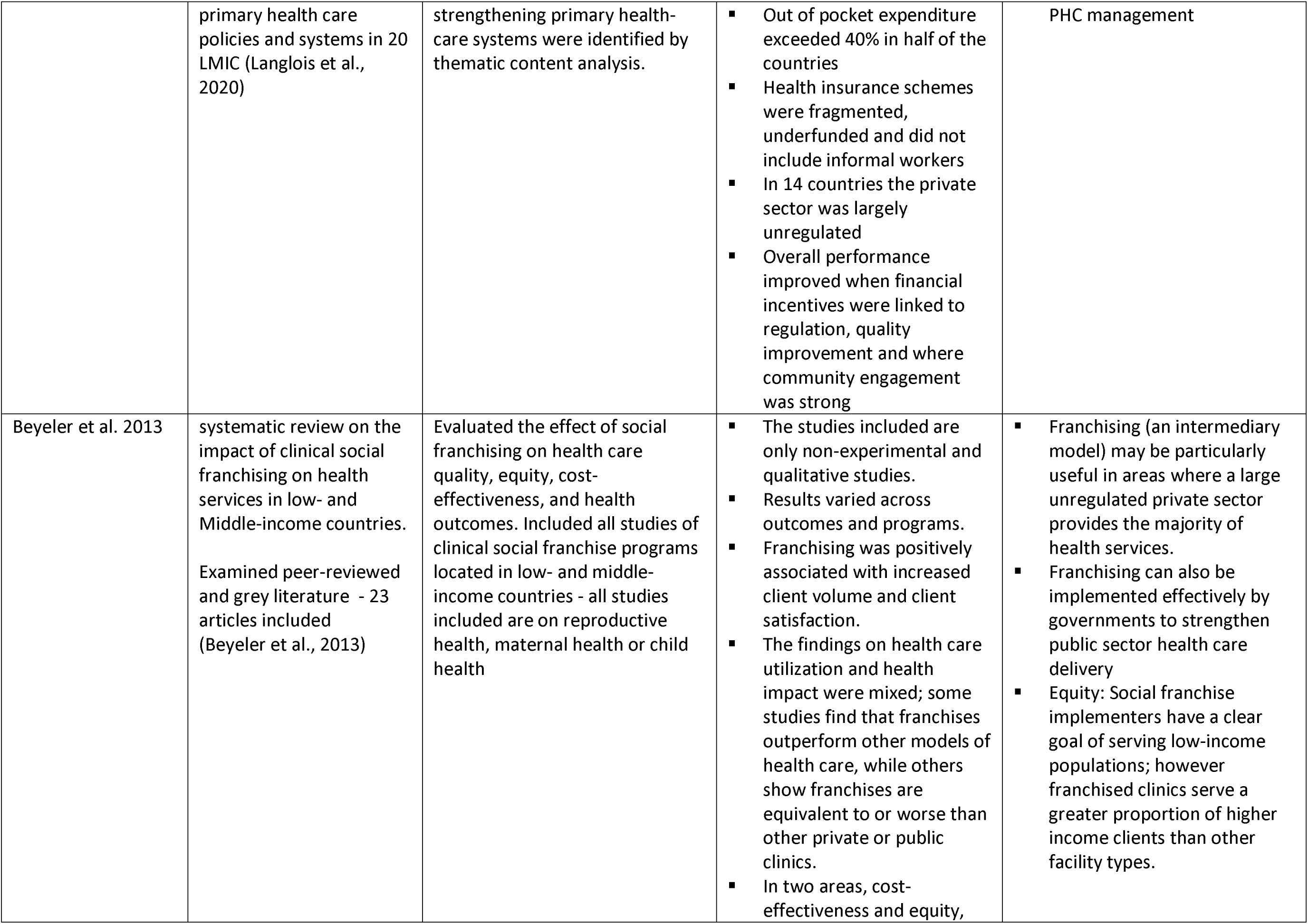
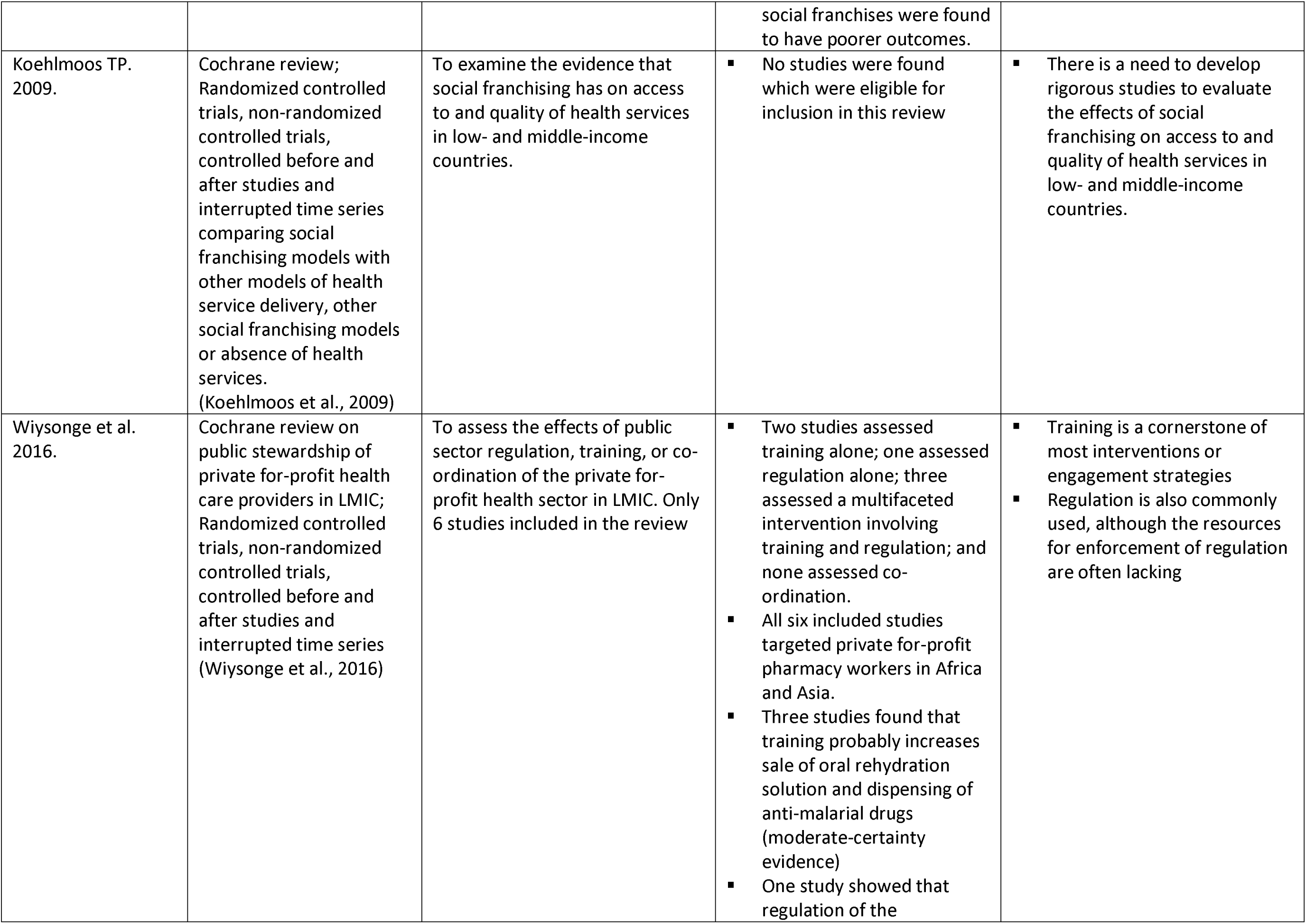
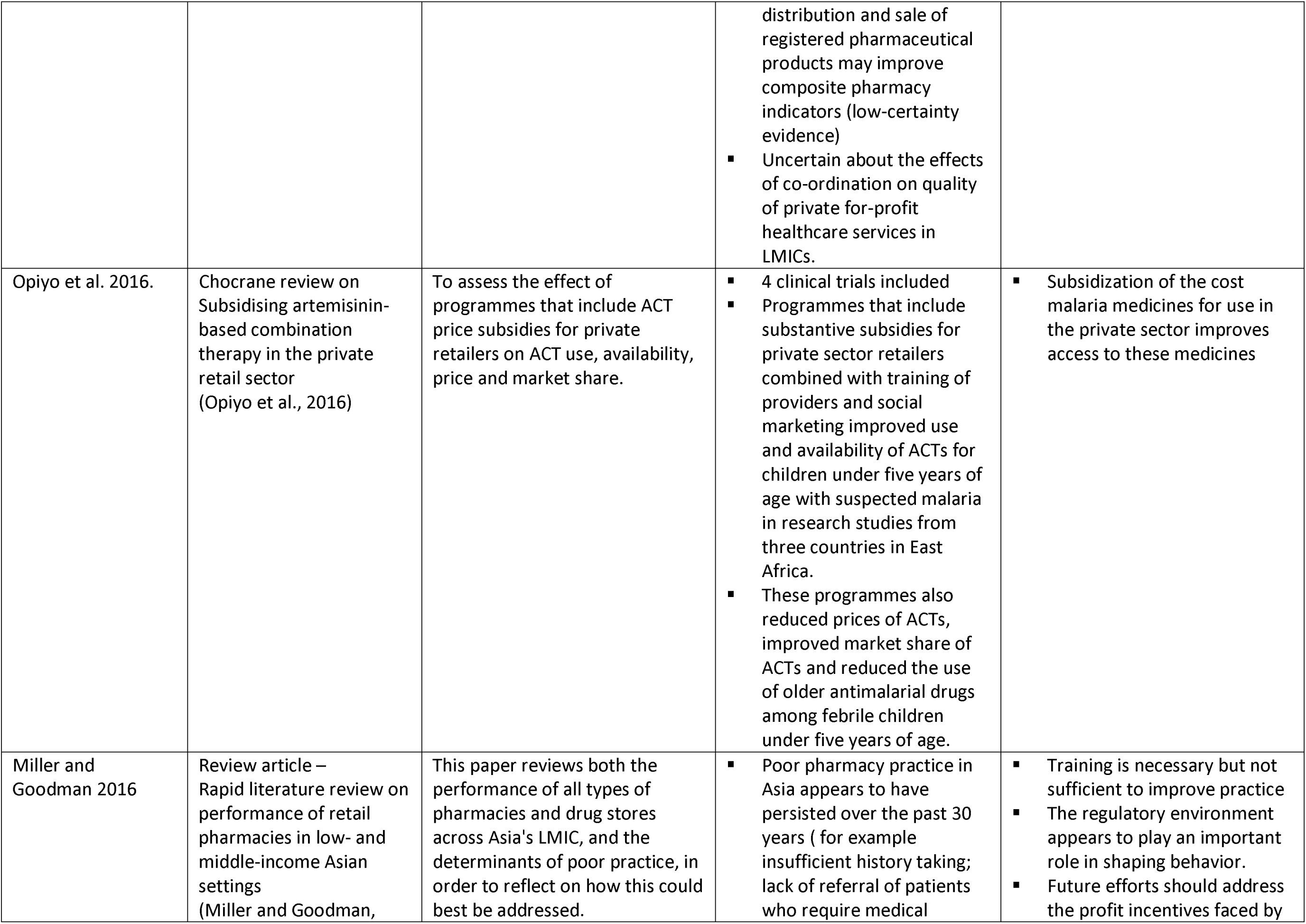
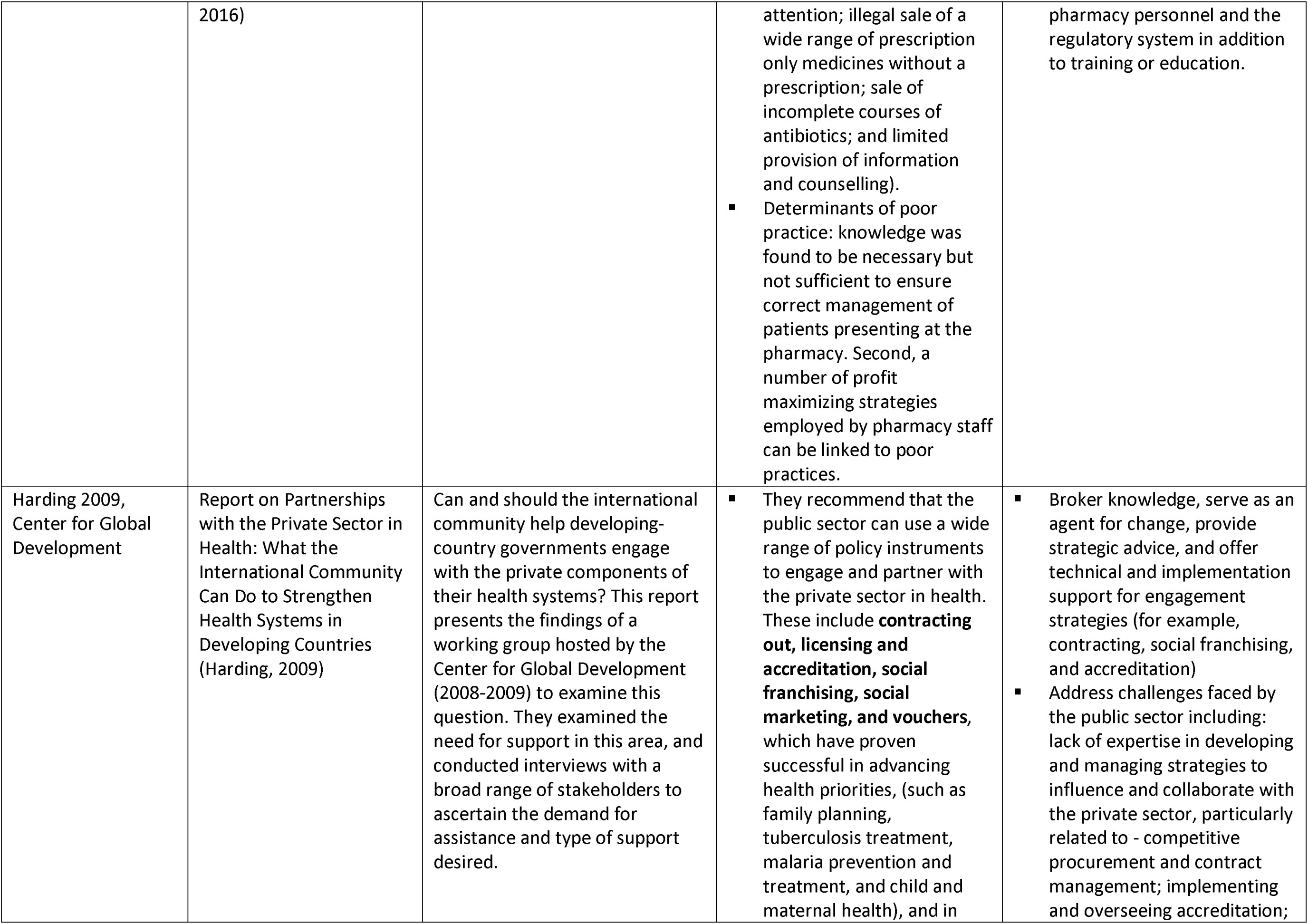
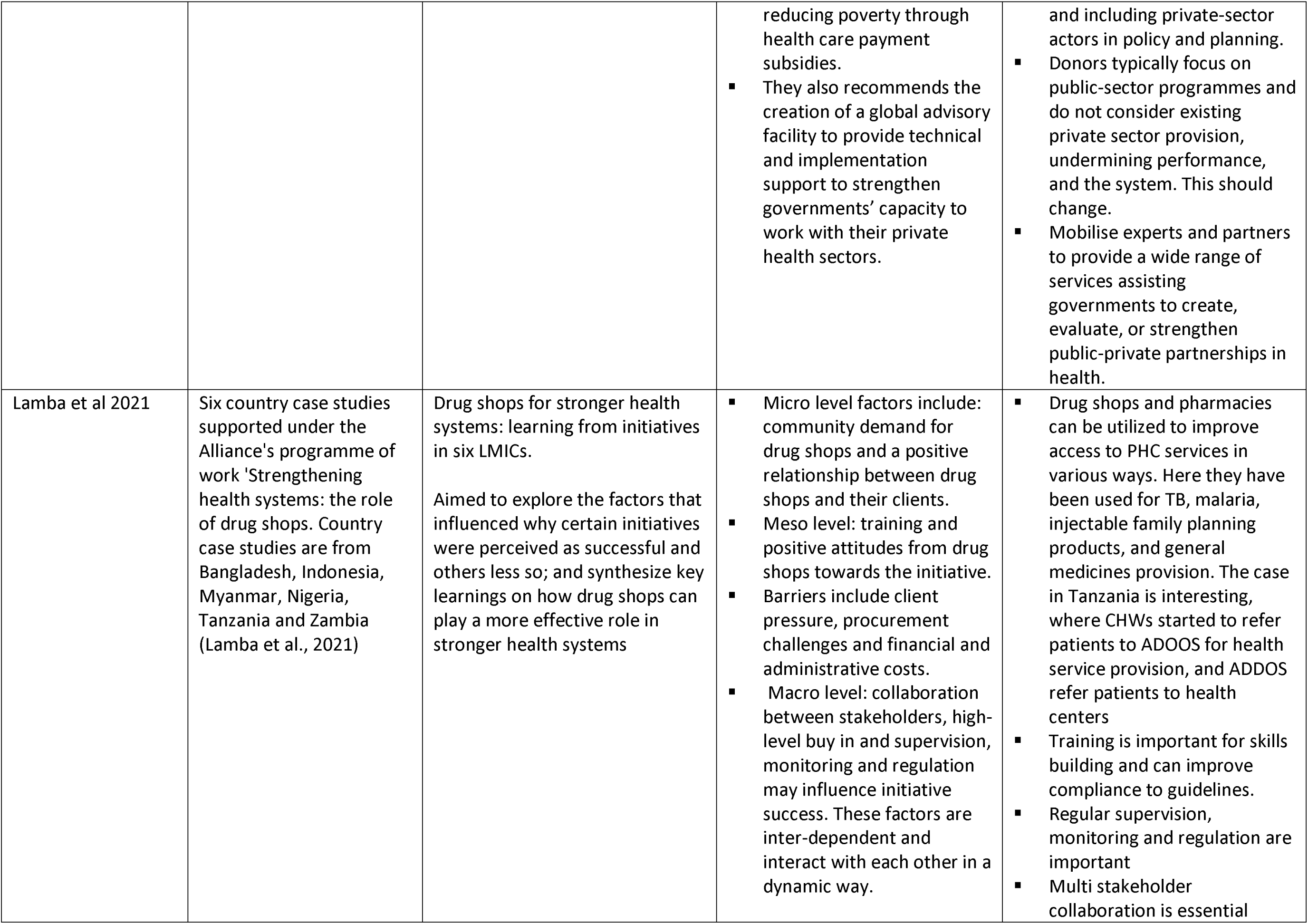
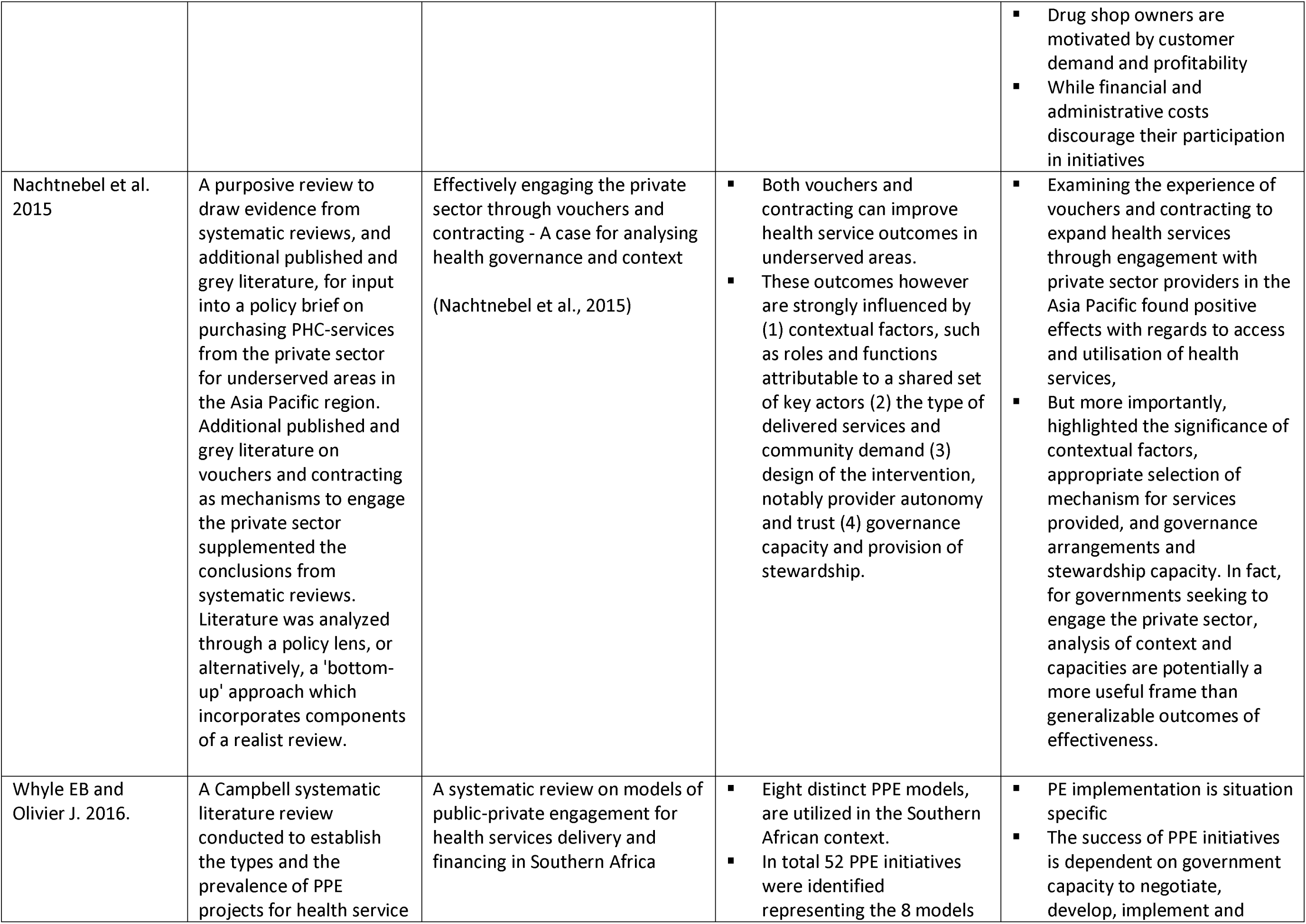
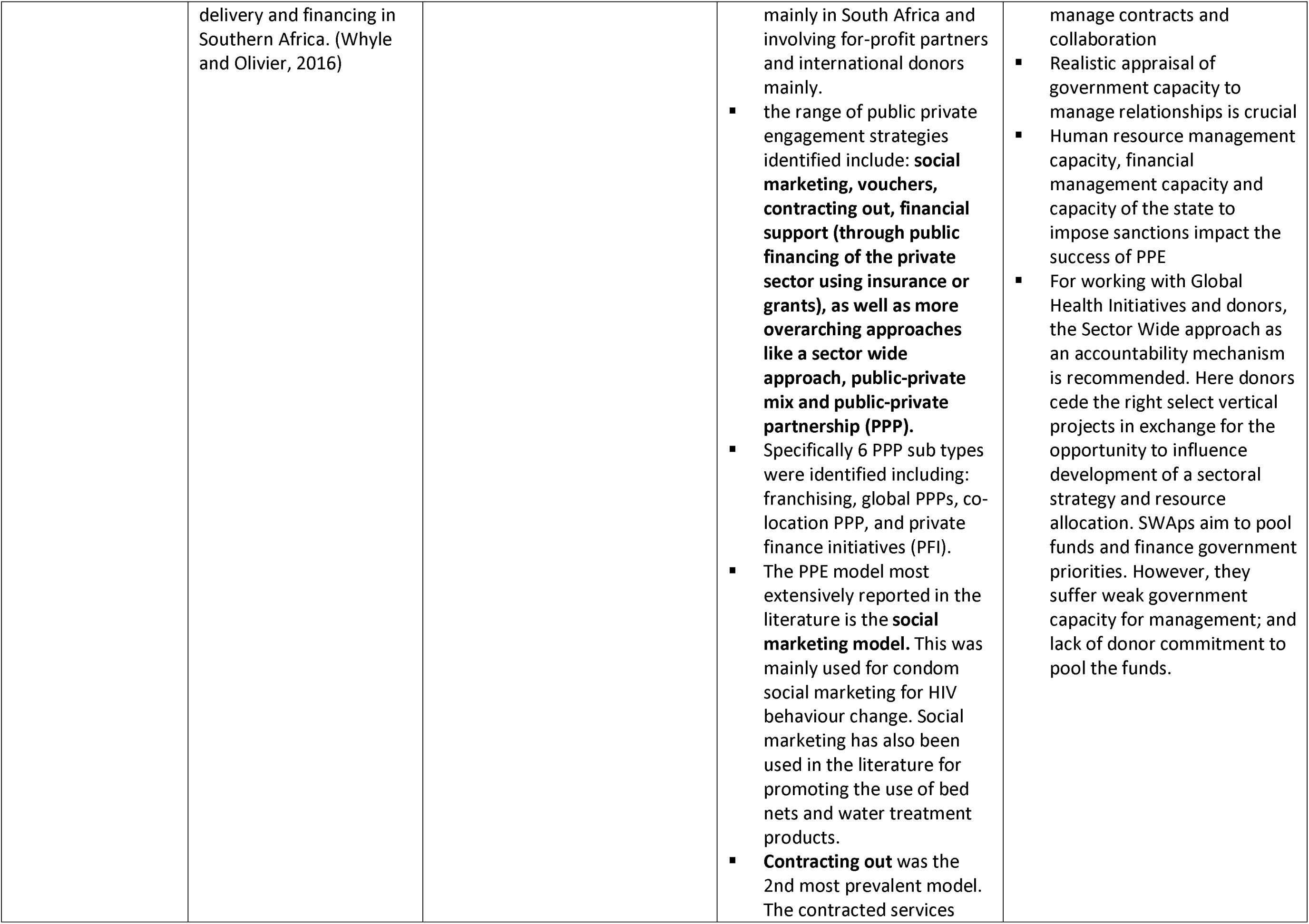
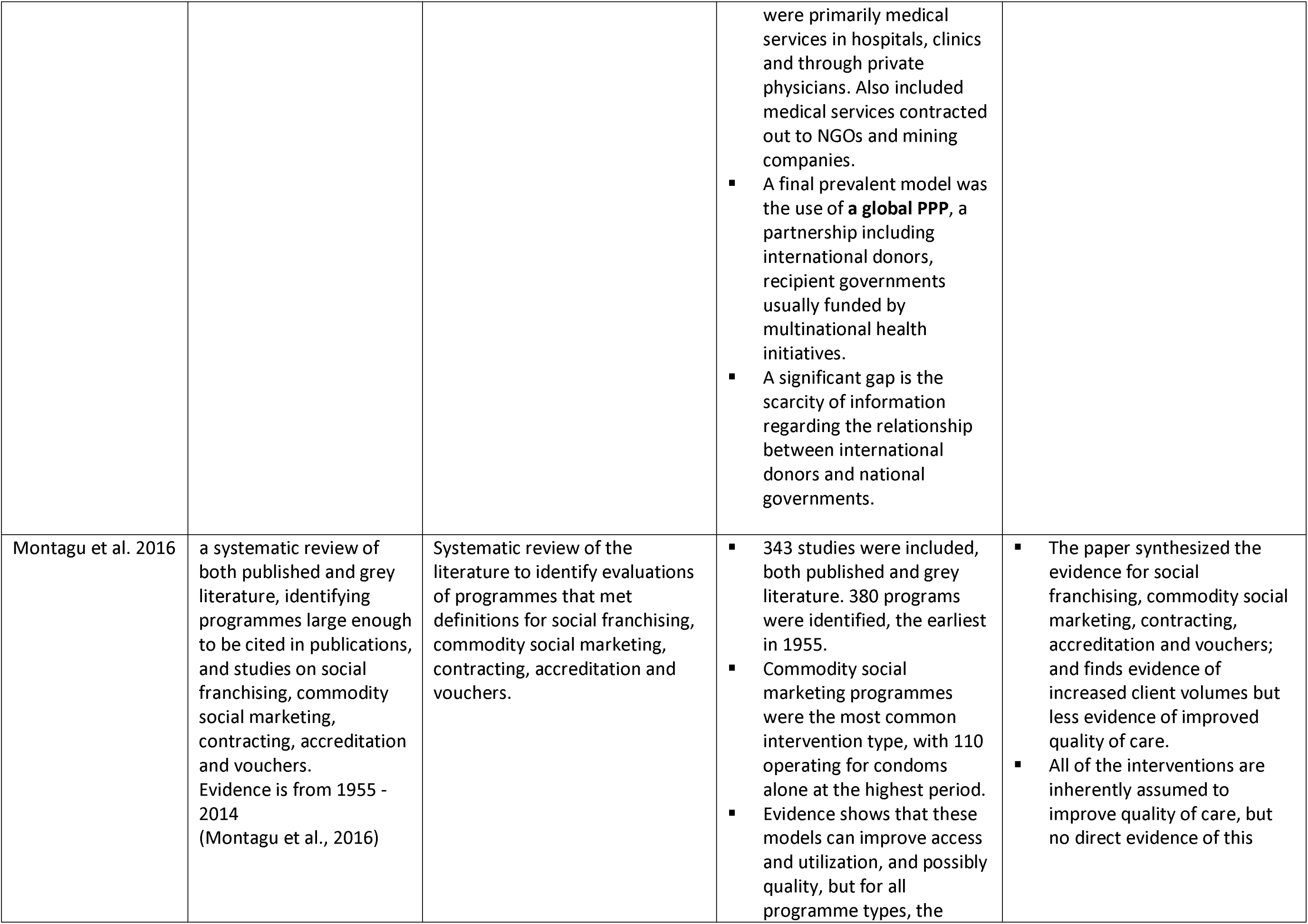
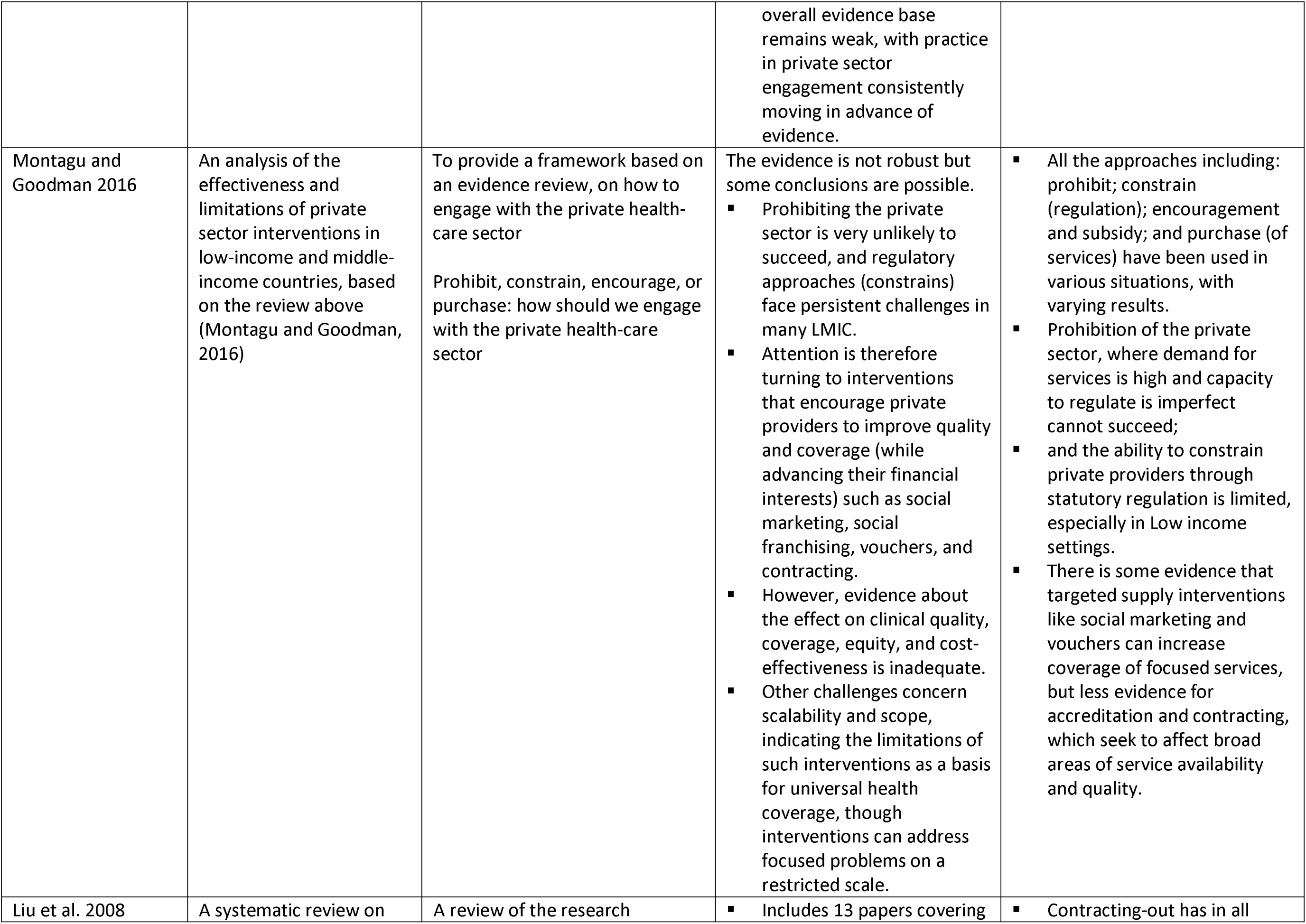
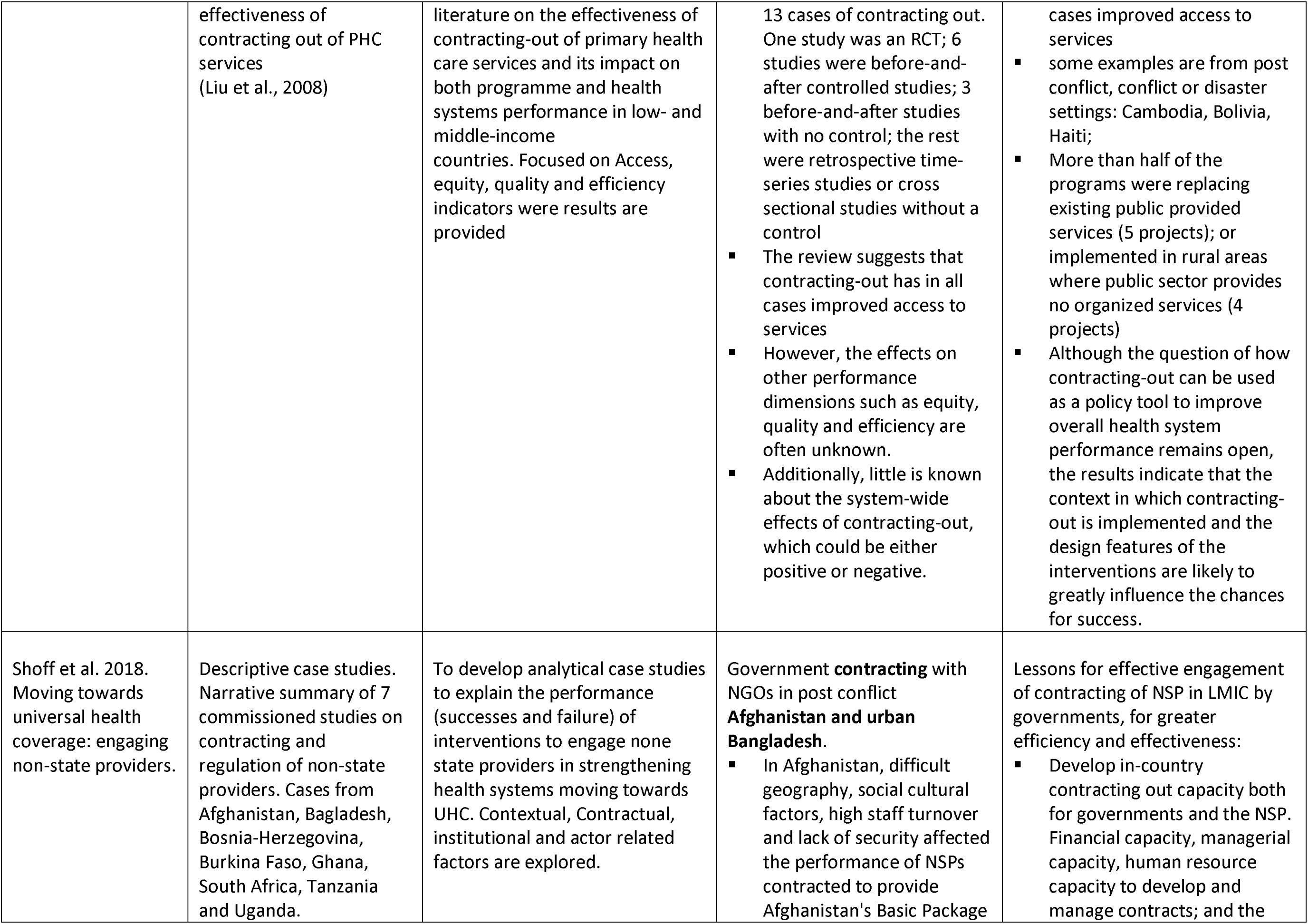
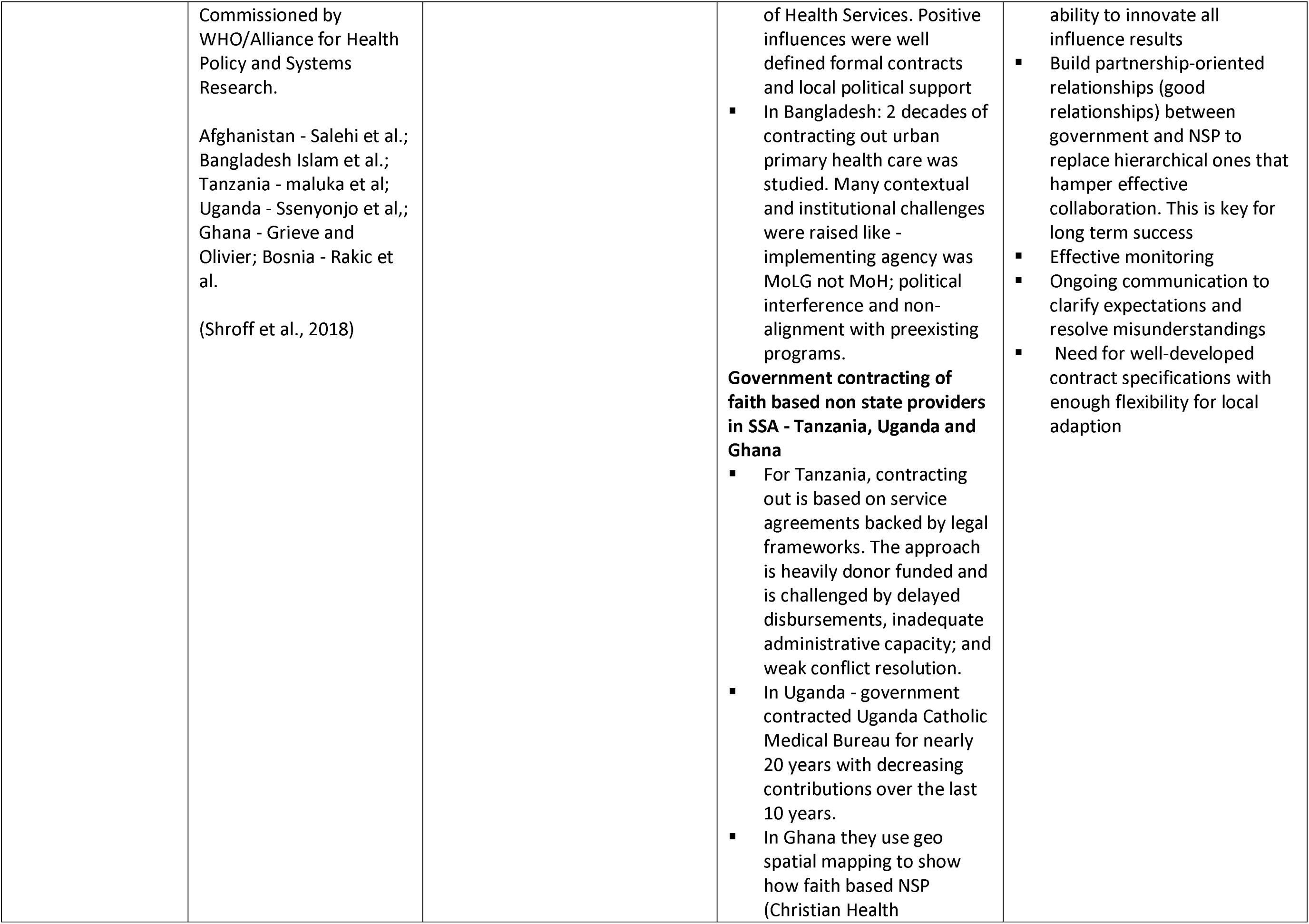
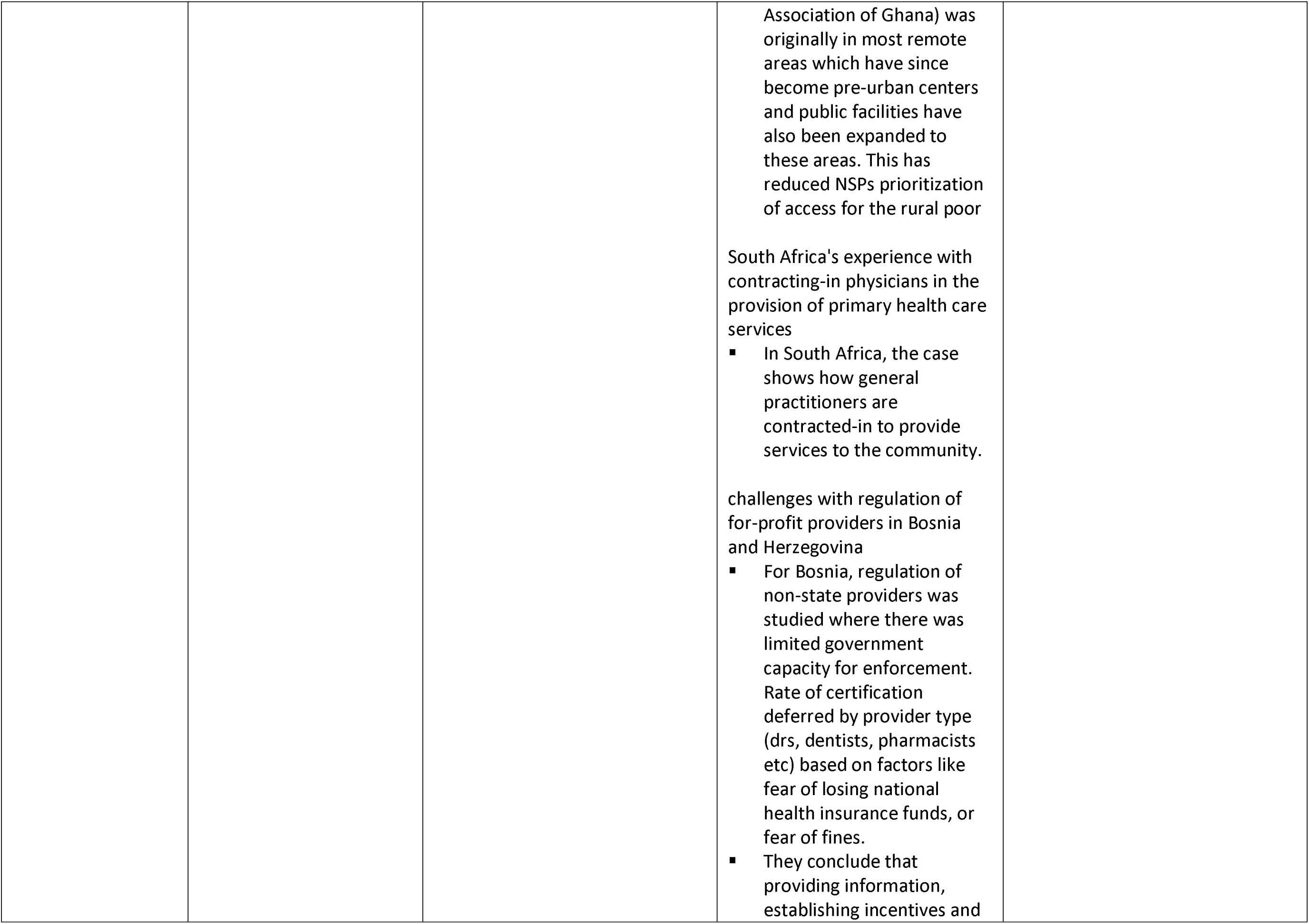
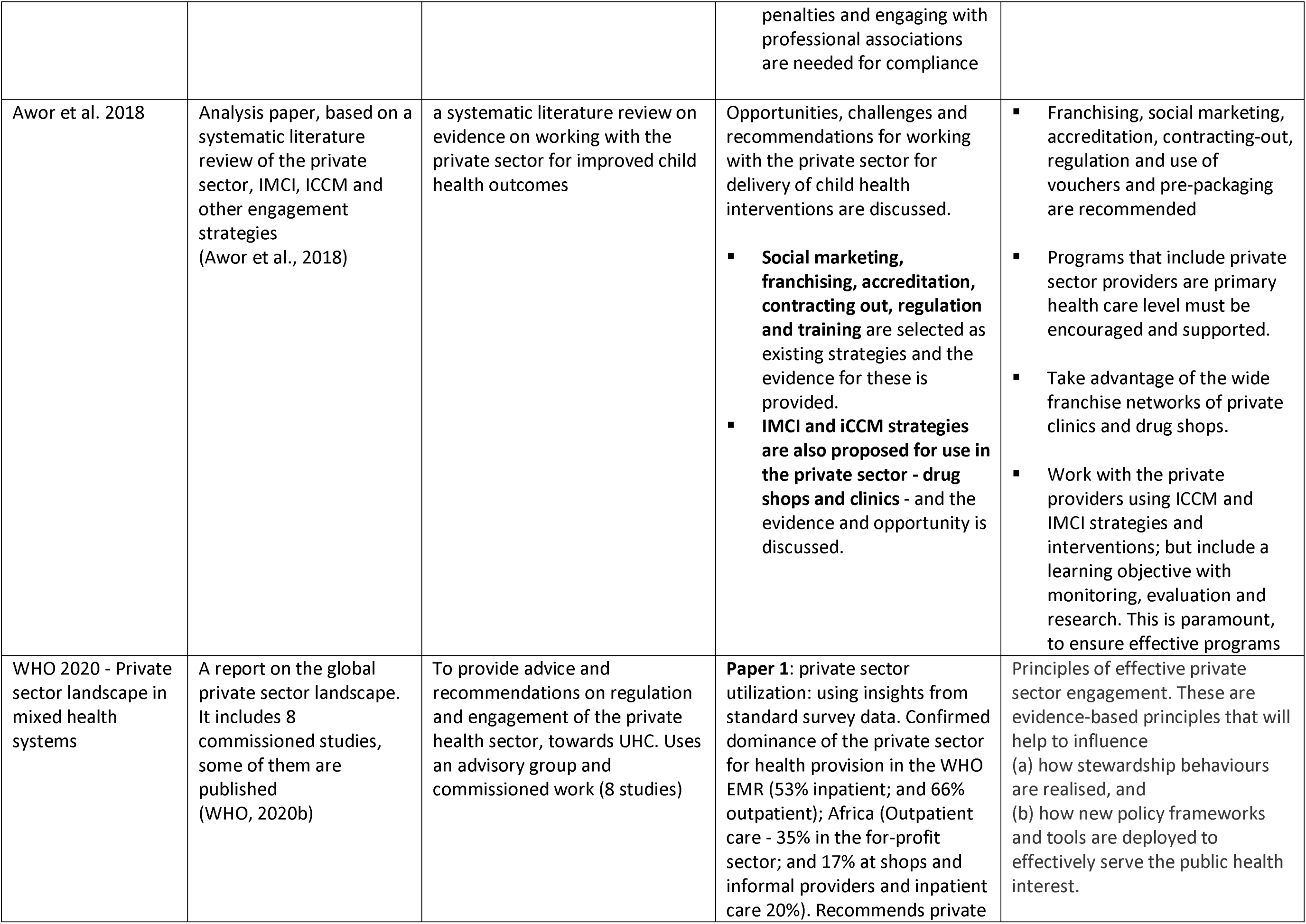
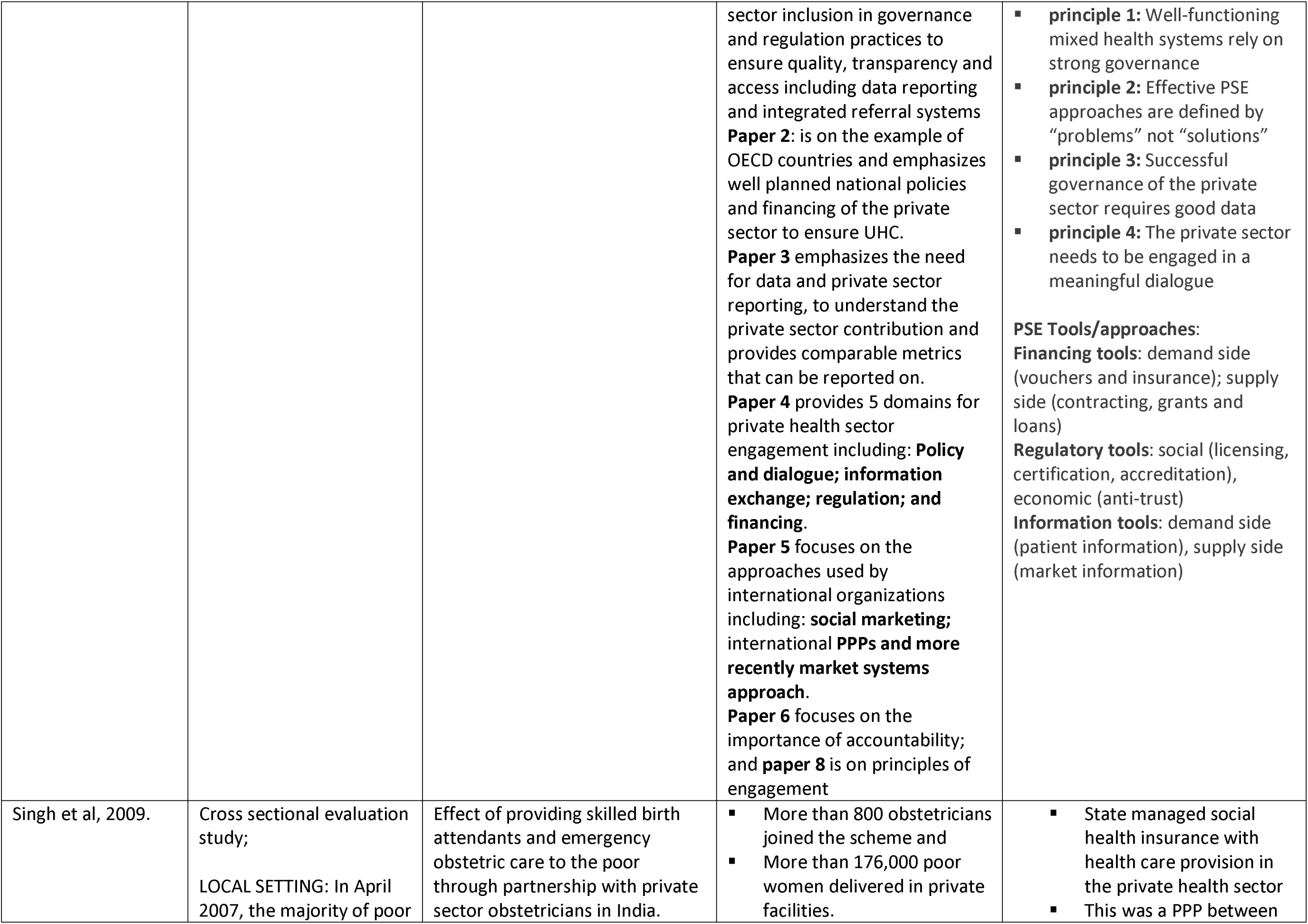
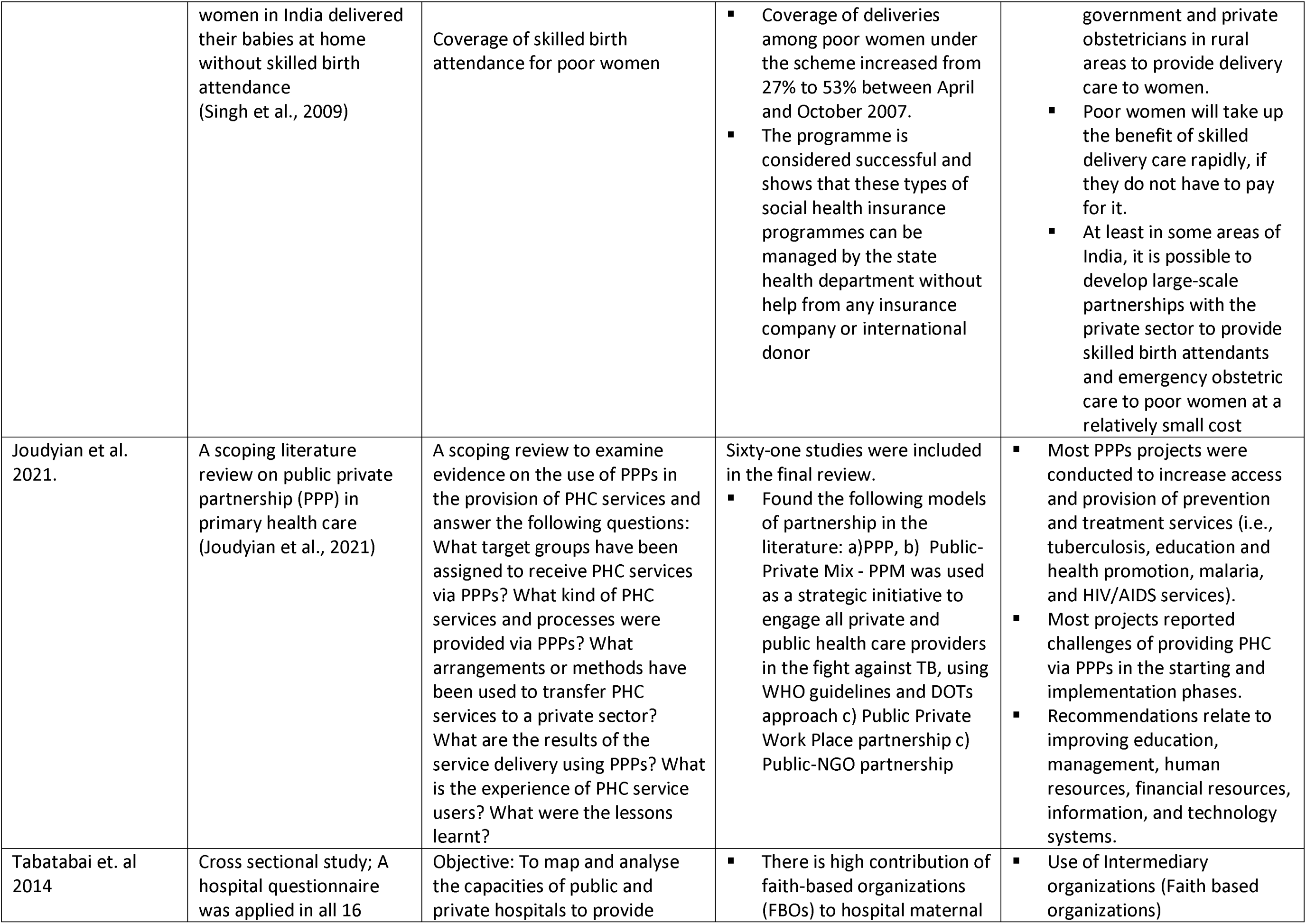
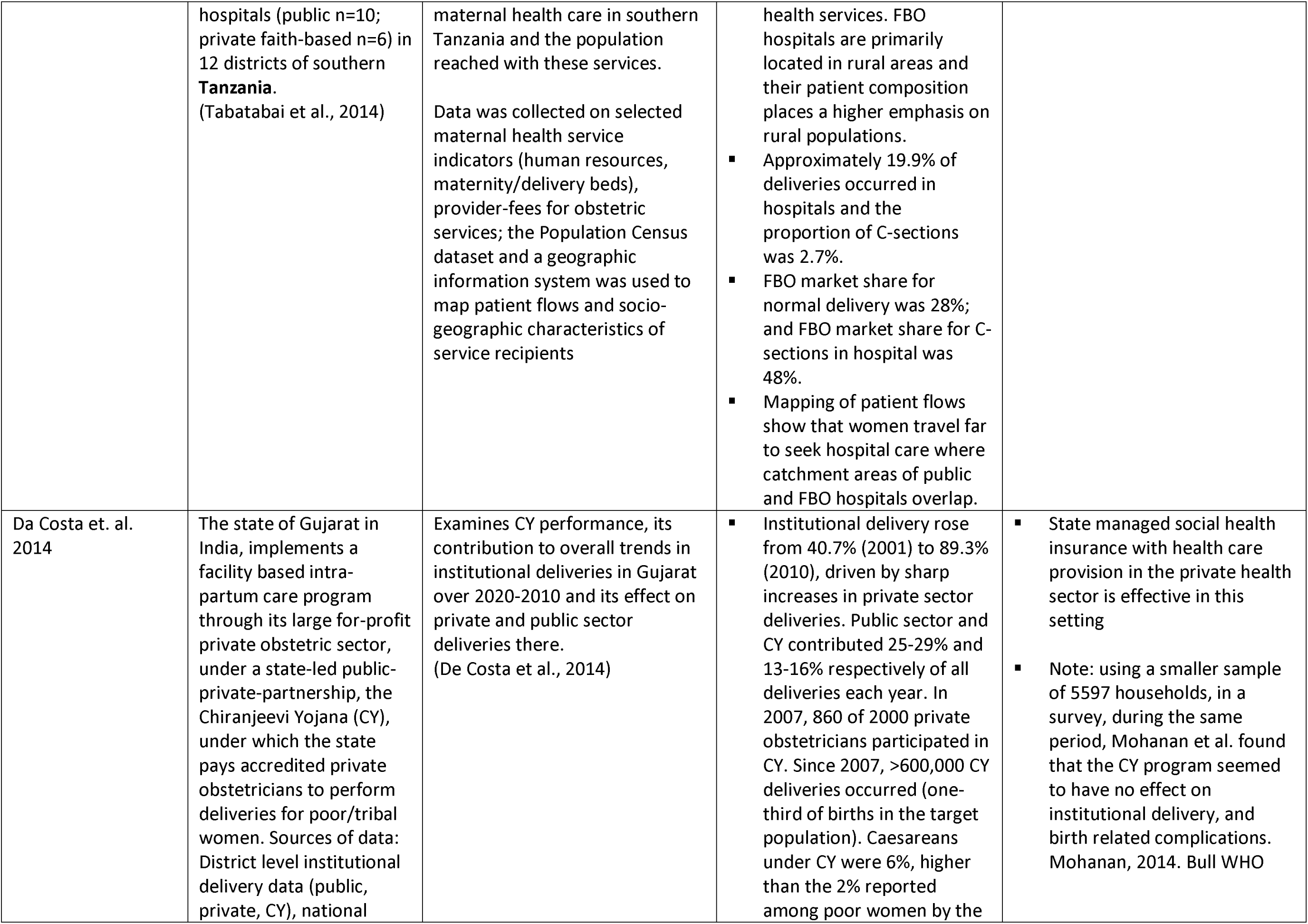
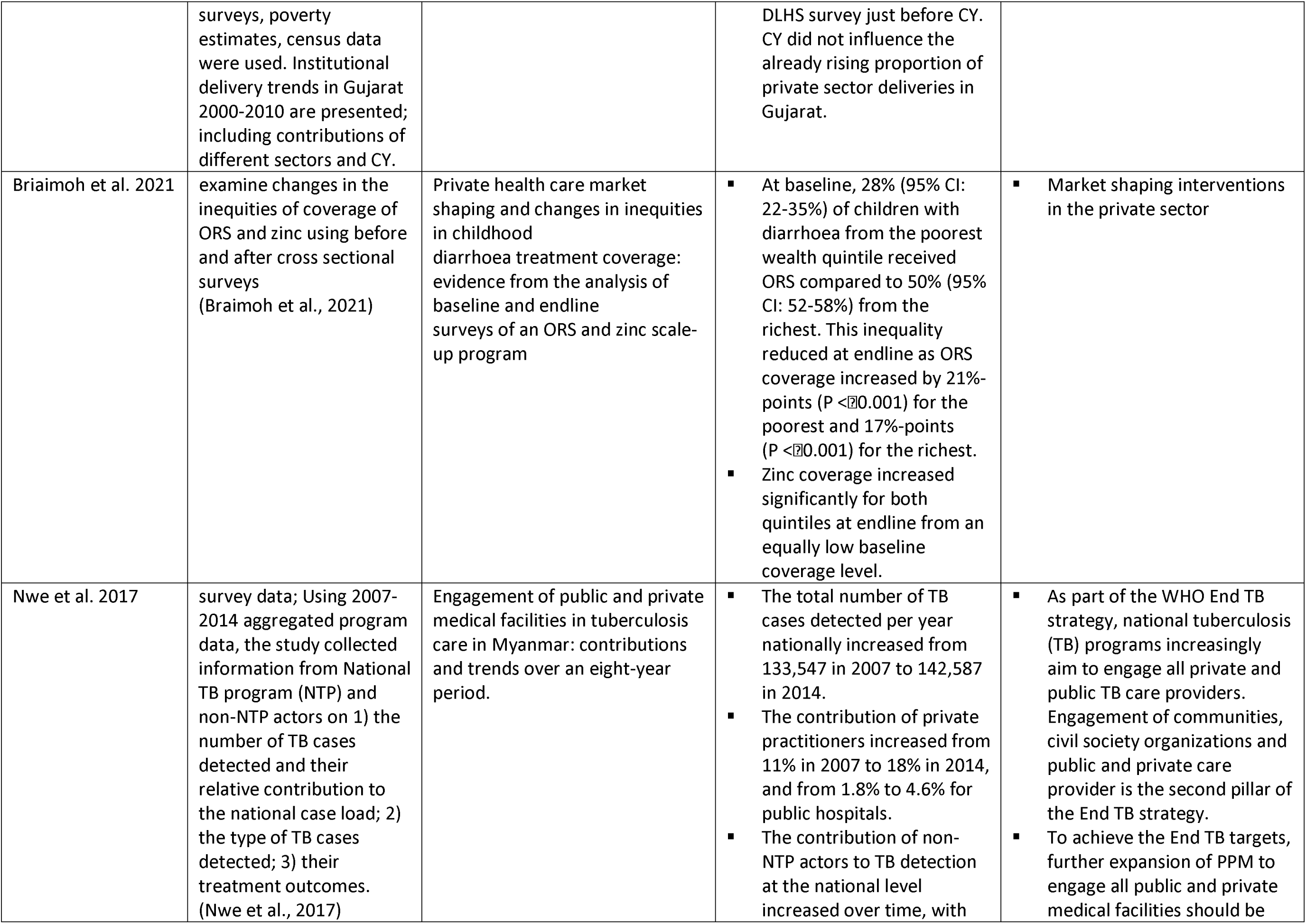
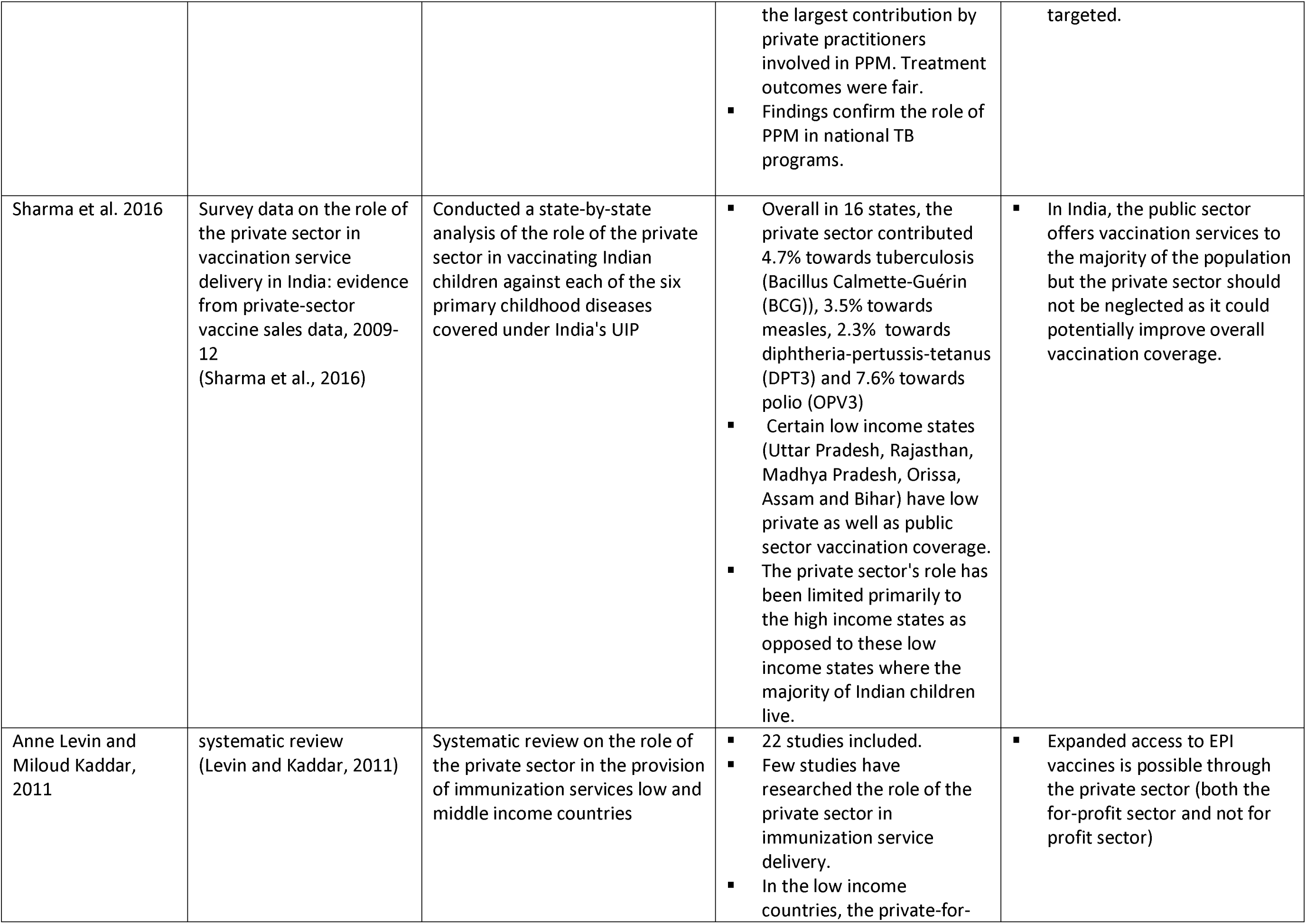
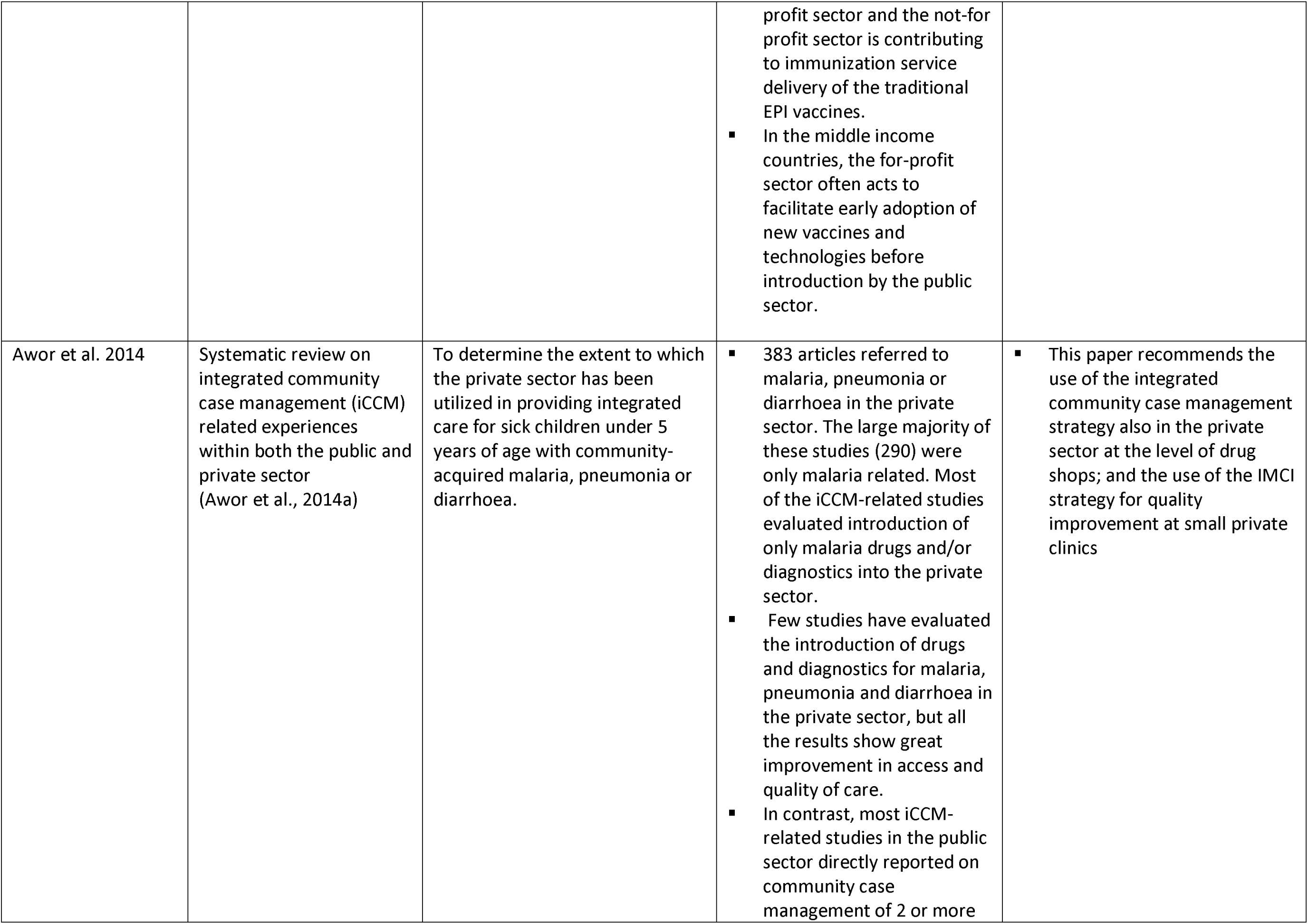
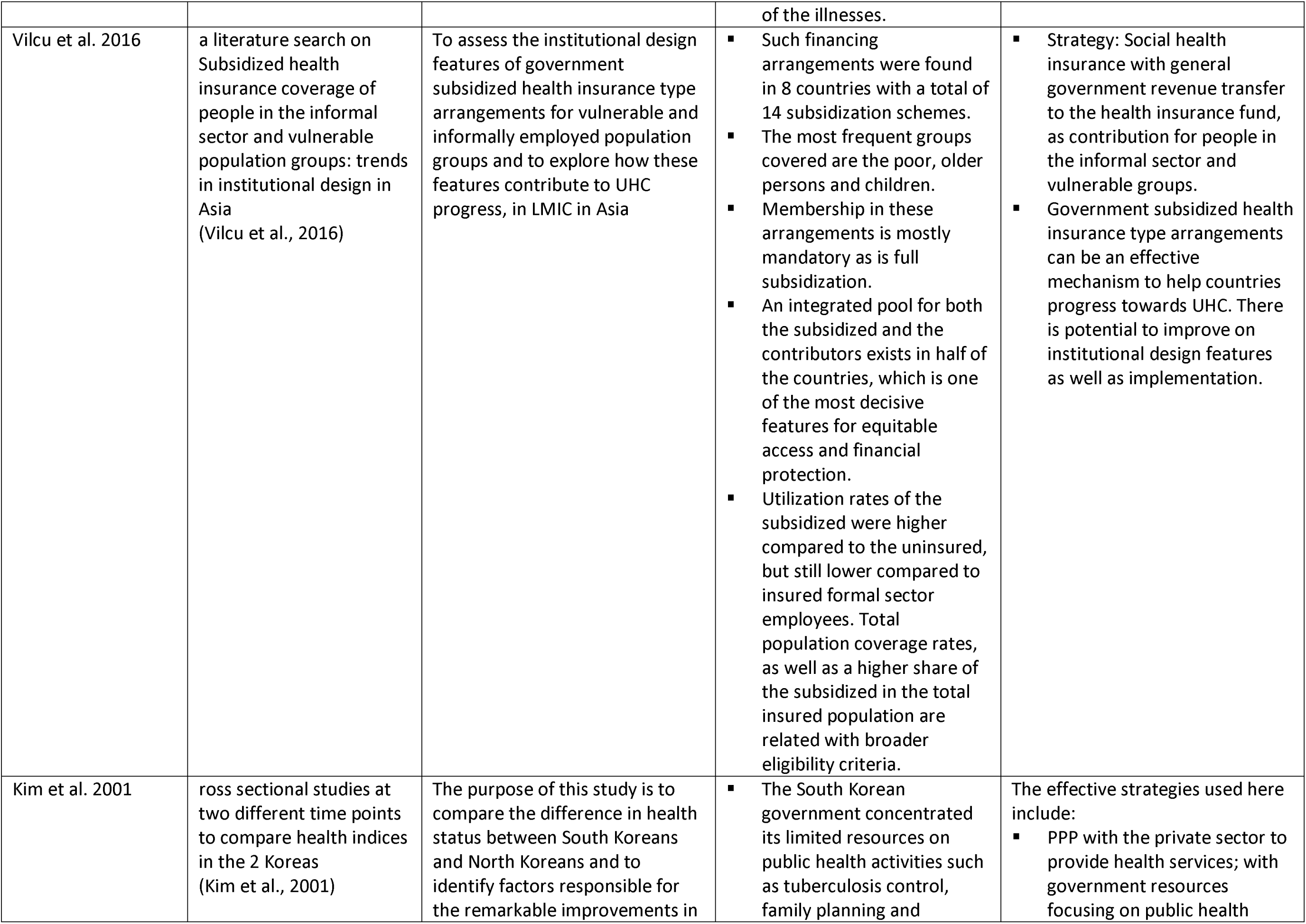
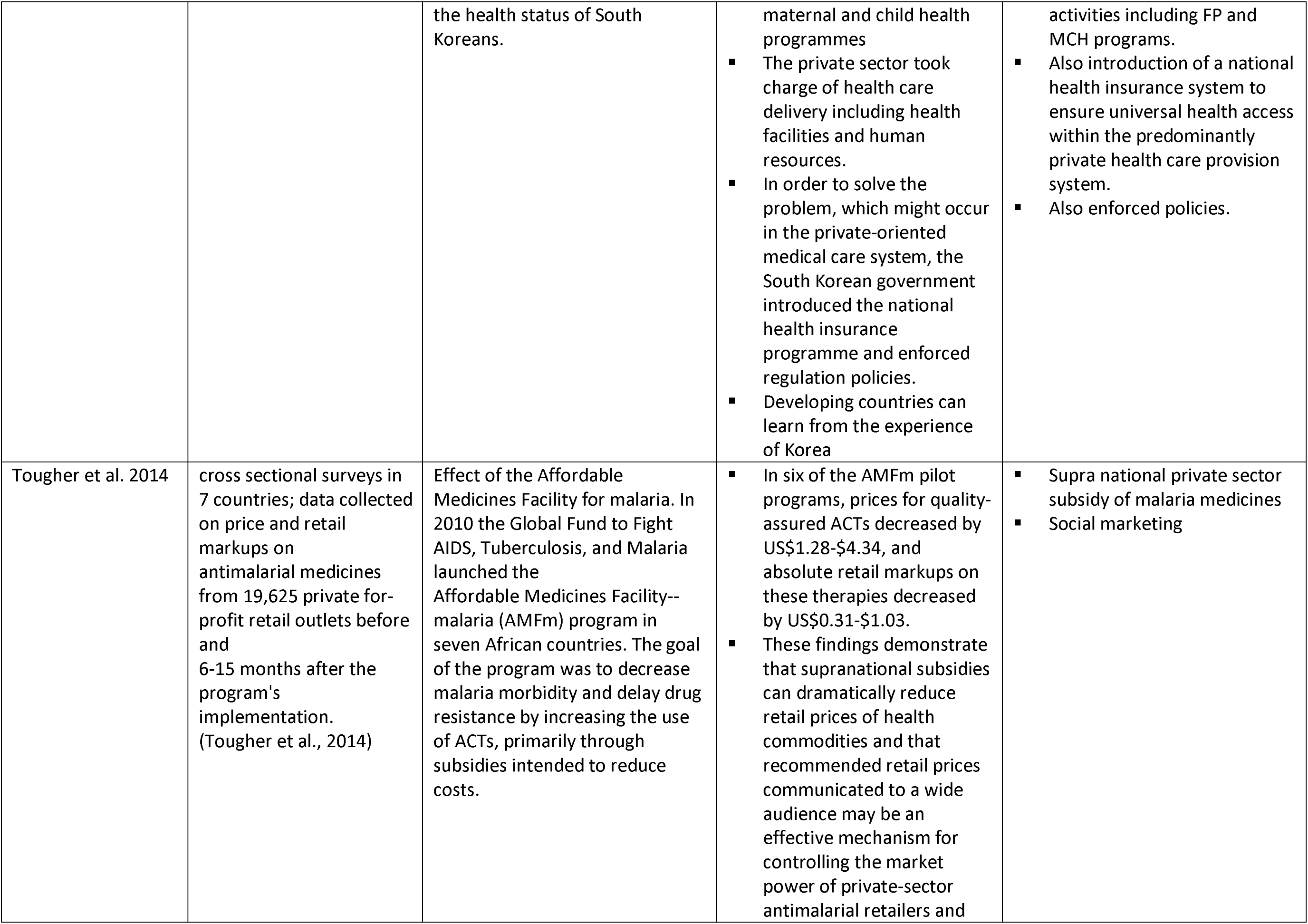
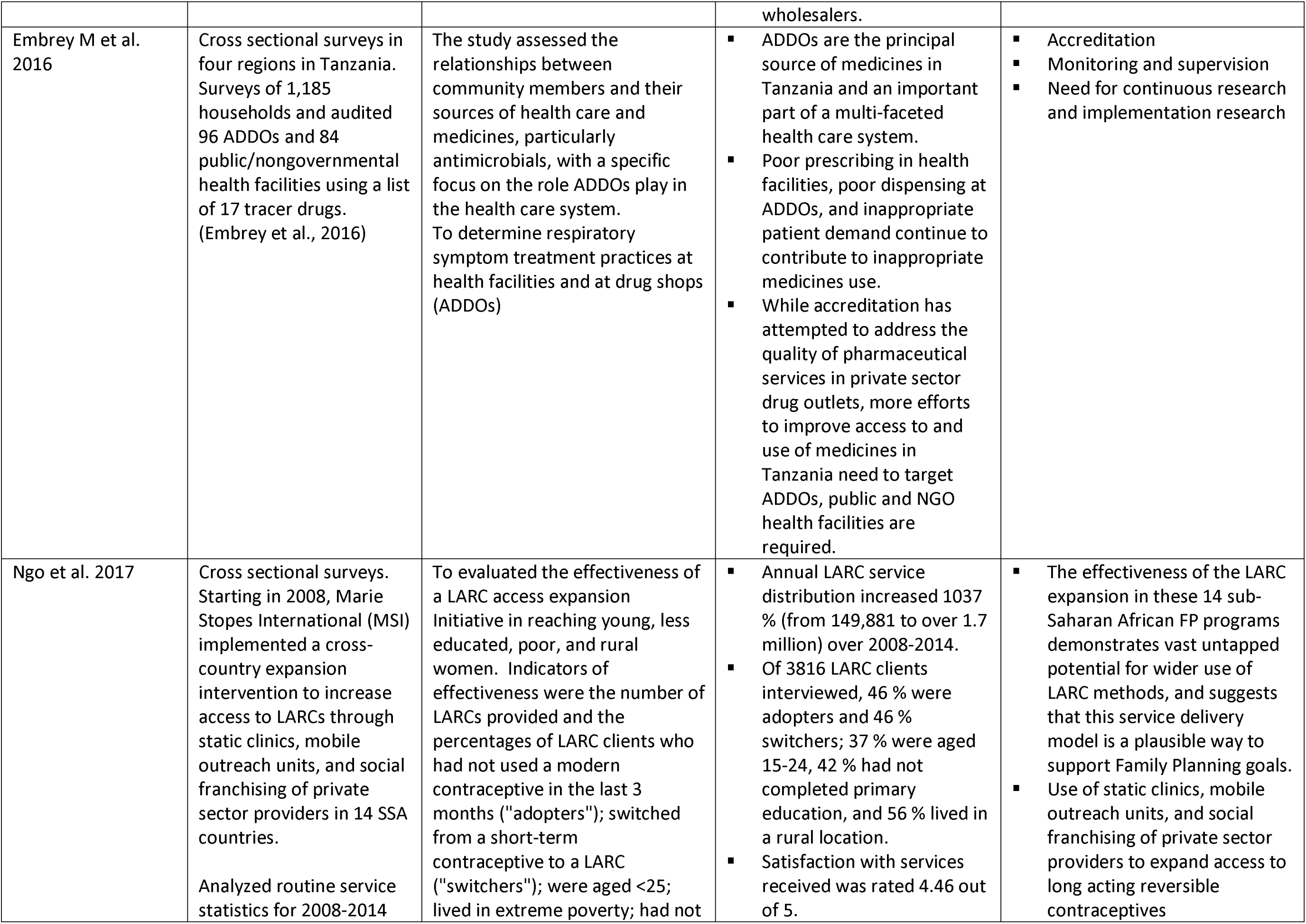
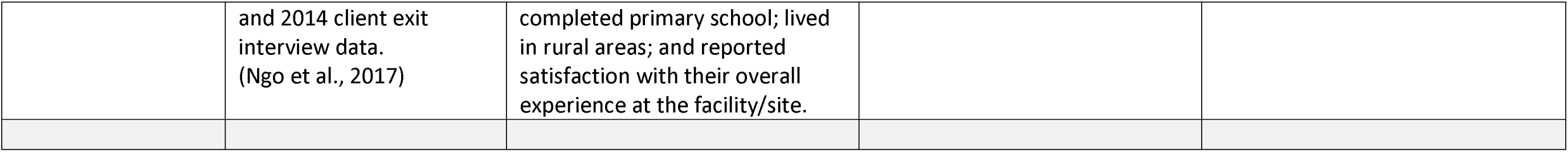
strategies for engagement with private health providers for maternal and child health primary health care service provision

## RESULTS AND DISCUSSION

A total of 3893 titles and their abstracts were identified and reviewed. After the abstract review, 332 of articles were taken to the next stage of full text review. Additionally, the references of the full text articles were scrutinized for relevant articles, yielding 60 more papers for full text review. Another 15 papers were identified from the grey literature for full text review. After the full review, 59** articles are included in our analysis and report.

### Extent of utilization of private providers for maternal and child health services in LMIC

Our findings confirm the dominant use of private health providers for outpatient care seeking, and substantial utilization of these providers for inpatient services including child birth. **Table 1** shows the studies included, and the extent of utilization of private health providers for specific maternal and child health services including family planning services, antenatal care, delivery services, and management of fever, malaria and pneumonia in children. Most of this data is multi country data, aggregated from Demographic Health Surveys (DHS) between 2010 and 2019; and a few health facility surveys.

Using both DHS and Multiple Indicator Survey data from 65 LMIC between 2010 and 2019, Montagu and Chakraborty showed that the private sector provides nearly 40% of all health care in the WHO PAHO, AFRO and WPRO regions, as well as 57% in the SEARO and 62% in the EMRO regions. They also showed that public and private care vary less by wealth levels for outpatient care. For inpatient care, this is predominantly used by the wealthy (Montagu and Chakraborty, 2021). Similarly, Grepin et al., 2016 showed using aggregate data from 70 DHS surveys in LMIC that the private sector was the dominant source of care for diarrhea and fever or cough treatment for sick children (67 percent and 63 percent, respectively); and institutional delivery - 38%; ANC - 30%; Modern contraceptives - 39% (Grepin, 2016). These findings are quite similar between all the other results from DHS surveys as in table one, irrespective of the period of review, that about 20-40% of deliveries occur in the private health sector (Benova et al., 2015, Benova et al., 2017, Dennis et al., 2018, Brugha and Pritze-Aliassime, 2003, Joe et al., 2018) and 30-50% of all family planning commodities are obtained from private health providers (Campbell et al., 2016, Dennis et al., 2018, Grepin, 2016).

While the poor and wealthy equally seek outpatient care from private health providers (Montagu and Chakraborty, 2021), this is not the same for inpatient care, which is predominantly used by the wealthy in the private health sector (Limwattananon et al., 2011, Montagu and Chakraborty, 2021). Generally, as wealth levels increase, private sector utilization increases for almost all services (Grepin, 2016, Brugha and Pritze-Aliassime, 2003, Campbell et al., 2016, Limwattananon et al., 2011, Waters et al., 2008).

Using world health survey data from 39 low and middle income countries, Saksena et al. 2012 determined the utilization rates, out of pocket payment (OOP) and per visit charges for outpatient and inpatient stays in both the public and private sector. They found that 65% of total OOP for inpatient services was paid at public health facilities and 45% of total OOP for outpatient services was also paid to public providers (Saksena et al., 2012). This high OOP payment at public facilities was mainly for the purchase of medicines (and less for consultation fees), showing problems with the existing policies in many LMIC where user fees were abolished at public health facilities.

### Effective private provider engagement strategies for maternal and child health services at the primary health care level

Generally, private health providers are engaged by the public sector using two overarching approaches: either through voluntary public-private partnerships, or through mandatory approaches for example licensing of private providers/facilities or mandatory reporting of notifiable diseases, particularly the case of tuberculosis diagnosis, treatment and reporting (using the Global TB program public-private mix approach). Various private sector engagement strategies and tools exist in the literature. Based on key literature, the consolidated scientific evidence on the strategies is presented in table 2, as well as recommendations on how to effectively utilize the specific strategy. The table also provides details on the level of success with different strategies.

The results in table 2 are mainly from published papers, particularly review papers on specific private sector engagement approaches; additional individual peer reviewed studies; as well as a few key reports from agencies and organizations that specialize in private health sector work. The literature congregates towards **macro** level (national) **meso** (intermediary level) and **micro** (individual level) **influences on private health providers**. The key strategies for private provider engagement identified through this review can be grouped under: **financing strategies, regulatory strategies, and policy** (including policy development) **strategies**.

The **Financing strategies** particularly include supply side strategies like contracting out, grants and loans as well as demand side strategies like vouchers and (social) health insurance. The **regulatory strategies** include licensing and accreditation which are the most common, as well as the less implemented but extremely effective strategies of monitoring and supervision, particularly support supervision. The **policy strategies** include existence of policy and/or guidelines for public private engagement, participatory policy development and effective dialogue mechanisms. **Figure 1** provides a summary of the effective strategies for engaging the private sector for maternal and child health service provision, based on our evidence review and synthesis.

**Figure 1:**
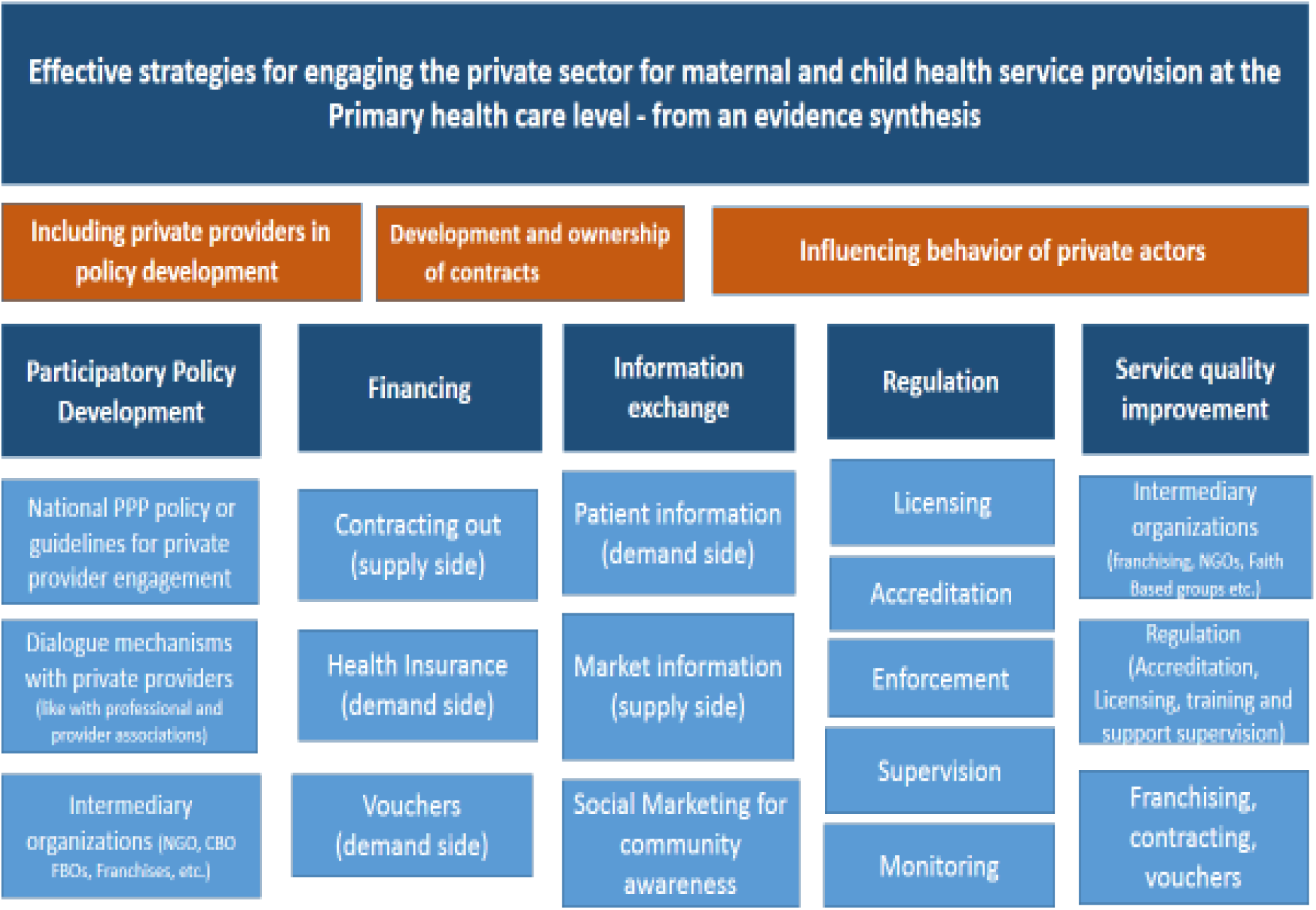
Effective strategies for engaging the private sector for maternal and child health service provision

**Intermediary organizations** are a dominant model utilized for coordination and engagement of private health providers. These are organizations that form networks between small-scale private providers in order to interact with governments, patients, and vendors while performing key health systems functions that are challenging for individual private providers to do on their own. Many and diverse examples of intermediary organizations exist in the literature, and the key ones identified are social franchising networks, non-governmental organization (NGOs) and faith based organizations (FBOs) which typically create strong networks of private health care providers for service quality improvement, regulatory compliance and provider representation (R4D, 2016).

The importance of **data** for understanding the private sector, its users (patients) and suppliers (providers) as well as the contextual and regulatory environment in which they operate cannot be over emphasized. Data allows policy makers to understand the private health sector, its contribution to national health goals and opportunities for better collaboration. **Information exchange** between the providers and regulators is therefore a key requirement for effective engagement. Additionally, provision of information to communities in order to empower them to make correct health choices and purchase quality health products is important. This has been done extensively through commodity social marketing (for example for purchase of bed nets or condoms), which is the commonest approach that has been used to engage private providers to sell quality products. Details of all these strategies is provided further below.

As in the figure above, engagement with private health care providers for effective service delivery should be undertaken at all the system levels including the macro level, meso and micro levels. At the macro (national) where policy development and decisions are made, these policy guidelines should be developed in collaboration with private providers and an attempt should be made to include the diverse groups of private health providers through the intermediary organizations that they subscribe to (NGOs, FBOs, Franchise organizations etc.); or through their direct representatives for example health professional bodies. Clear mechanisms for dialogue between the providers, regulators and decision makers is important.

At the Meso (regional and district) level, it is important to again involve the private providers in the decision making and particularly negotiation and ownership and implementation of contracts and other financial tools that are available in the system. At the individual or micro level, various options are available for influencing the provider behaviour and service quality. These include strict enforcement of regulatory tools for example licensing and accreditation of providers, as well as training and support supervision.

### Contracting out

Contracting out is a purchasing mechanism used to acquire specified services, of defined quality, at an agreed price, from a specific private provider and for a specific period of time. Governments may purchase clinical or non-clinical services from private providers to complement public provision. Contracting out has been shown to be effective at increasing access and use of health services, and in reducing out of pocket health expenditure in conflict or fragile states as well as in stable environments (Liu et al., 2008, Lagarde and Palmer, 2009, Nachtnebel et al., 2015, Shroff et al., 2018). Contracting out is no better or no worse than government health-provided services (Odendaal WA, 2018). See table 2 for more detailed information based on the studies on contracting out.

### Social franchising

A franchise is a contractual arrangement between a health service provider and a franchise organisation, which aims to improve access to quality and price controlled services. Franchisees are trained in standardised practices for which prices are predefined, and they benefit from advertising the logo or franchise name. Franchising is associated with increased numbers of clients, patient satisfaction, physical accessibility, and improved quality (Beyeler et al., 2013). Further research is needed to elucidate the effect of franchising for quality, health impact, equity, cost effectiveness, and the value of franchising in other healthcare sectors like child health (Beyeler et al., 2013, Koehlmoos et al., 2009, Montagu et al., 2016).

Numerous social franchising programmes already exist around the world, providing an opportunity to expand access to care rapidly and to standardise and improve the quality of care. (Montagu et al., 2016, Awor et al., 2018, Viswanathan, 2014). This could form the basis for evaluation of private sector initiatives, provided that evaluation is built into further expansion of the social franchises.

### Regulation

Regulatory interventions are used to set up and ensure adequate technical quality of service providers. Regulation involves setting rules, sanctions, and ensuring adequate enforcement. Basic regulatory frameworks exist in most countries, particularly for pre-service training, registration and licensing requirements for health workers and premises (WHO, 2020b). Pharmaceutical market regulation aims to limit the availability of harmful drugs and unregistered products, minimise drug misuse, control the sale of specific drugs through prescriptions and regulate drug manufacture and importation. Regulation has a crucial balancing role within the private sector, although, inadequate resources are typically allocated for monitoring and enforcing regulations (Wiysonge et al., 2016, Montagu and Goodman, 2016, Shroff et al., 2018, Lamba et al., 2021). Co-regulation with professional associations, civil society, and communities provides additional benefit. (Lamba et al., 2021, WHO, 2020a, WHO, 2020b). See table 2 for more detailed information on the studies and recommendations related to regulation.

### Drug shops as a provider of maternal and child health services

As shown in table 1, drug shops are a dominant provider of medicines and health care services for adults and children in LMIC, particularly anti-malarial medicines, fever medication and family planning commodities. This group of providers has been effectively engaged through various ways. Training, accreditation, supervision and monitoring are effective ways of improving knowledge and compliance to guidelines when working with drug shops for improved primary health care outcomes (Lamba et al., 2021, Awor et al., 2018, Embrey et al., 2016). Organizing these drug shops through intermediary organizations like franchise networks and accreditation bodies has also been effective (Embrey et al., 2016, PSI, 2016). Programs that include price subsidies for example for ACT commodities, ORS Zinc, other child health commodities and programs which have utilized the integrated community case management strategy at drug shops have also been shown to be extremely effective. (Opiyo et al., 2016, Tougher et al., 2012, Tougher et al., 2014, Awor et al., 2018, Awor et al., 2014b, Braimoh et al., 2021). See table 2 for detailed information on studies and recommendations on utilizing drug shops for stronger health systems.

### Social health insurance and maternal and child health service provision in the private sector

Through this review we identified some papers on the role of state led and state funded social health insurance programs which expand access to maternal and child health services through the private sector. The evidence provided is from 8 countries in Asia based on a review, and two additional papers were from India and another paper comparing north and South Korea PHC interventions. Inclusion or private obstetricians and private health providers in public private partnerships for provision of child birth and other health services to people with social health insurance cover has been shown to greatly increase access to health care at the PHC level. (De Costa et al., 2014, Kim et al., 2001). Government subsidized health insurance type arrangements can be effective mechanism to help countries progress towards UHC, although there is room to improve the design of these schemes (Vilcu et al., 2016).

### The role of the private sector in the provision of immunization services

While the public sector offers vaccination services to the majority of the population in LMIC, there is some evidence on the role of the private sector in immunization service delivery. In the low income countries, the private-for-profit sector and the not-for profit (NGO) sector is contributing to expanded immunization service delivery of the traditional EPI vaccines. In the middle income countries, the for-profit sector often acts to facilitate early adoption of new vaccines and technologies before introduction by the public sector.(Sharma et al., 2016, Levin and Kaddar, 2011).

## CONCLUSIONS

Our results confirm that the private sector is the dominant provider of outpatient care for women and children in low and middle income countries, and a significant provider of reproductive and maternal health services including for inpatient care. More urban and wealthier women utilize the private sector for maternal health services, but equally poor and wealthier families seek out patient care for children in the private sector.

The Private sector plays different roles in different settings/countries and so there is no single intervention or approach which will work in all contexts. It is therefore important to utilize existing data to understand an individual context, the actors and relevant approaches for engagement.

Effective private health provider and private sector engagement must include the following dimensions and underlying strategies: a) private sector participation in public health policy development including development and ownership of contracting arrangements; b) and influencing private sector behavior through regulatory and financing tools as well as direct quality improvement interventions.

## SUMMARY OF KEY RECOMMENDATIONS FOR PRIVATE PROVIDER ENGAGEMENT

A summary of the recommendations highlighted in Table 2 is provided here. These recommendations are for response to specific challenges of private provider engagement that were also identified through this review.

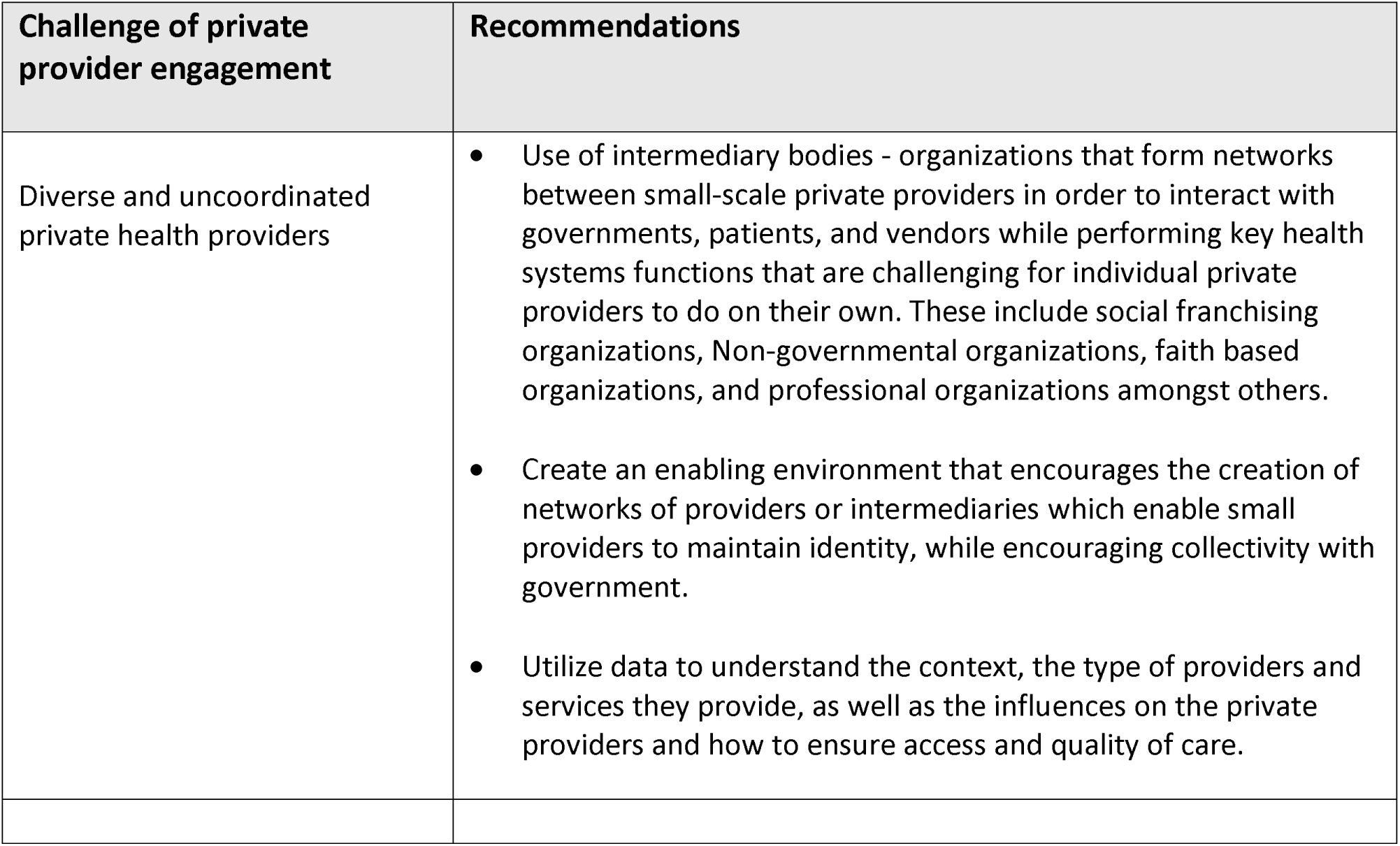
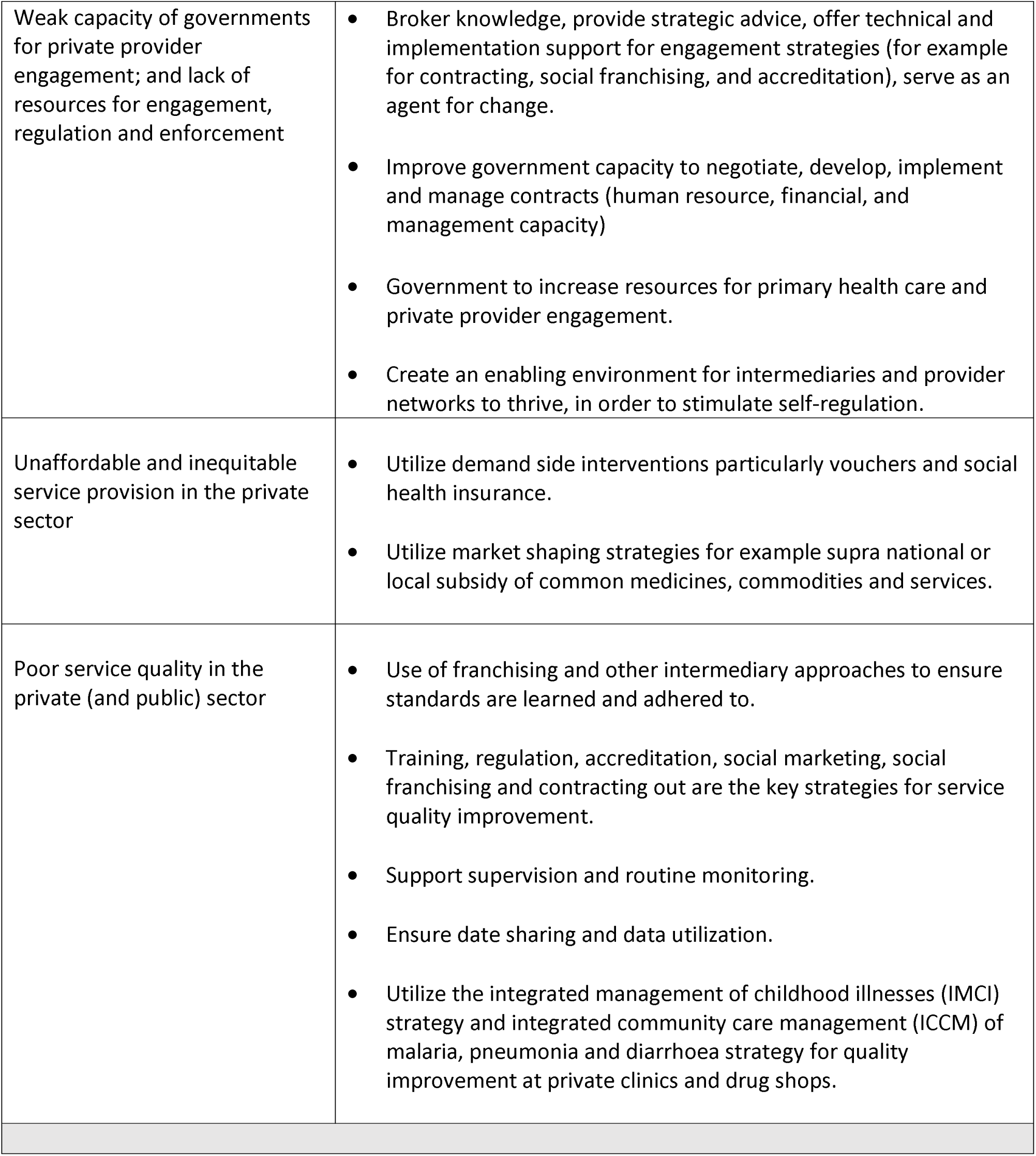

## Data Availability

All data produced in the present study are available upon reasonable request to the author

